# TACTILE EDGES AND MOTION VIA PATTERNED MICROSTIMULATION OF THE HUMAN CORTEX

**DOI:** 10.1101/2024.05.06.24306627

**Authors:** Giacomo Valle, Ali H. Alamari, Robin Lienkämper, John E. Downey, Anton R. Sobinov, Linnea J. Endsley, Dillan Prasad, Michael L. Boninger, Jennifer L. Collinger, Peter C. Warnke, Nicholas G. Hatsopoulos, Lee E. Miller, Robert A. Gaunt, Charles M. Greenspon, Sliman J. Bensmaia

**Affiliations:** Department of Organismal Biology and Anatomy, University of Chicago, Chicago, IL; Committee on Computational Neuroscience, University of Chicago, Chicago, IL; Rehab Neural Engineering Labs, University of Pittsburgh, Pittsburgh, PA; Department of Physical Medicine and Rehabilitation, University of Pittsburgh, Pittsburgh, PA; Department of Bioengineering, University of Pittsburgh, Pittsburgh, PA; Department of Neurological Surgery, University of Chicago, Chicago, IL; Neuroscience Institute, University of Chicago, Chicago, IL; Department of Neuroscience, Northwestern University, Chicago, IL; Department of Biomedical Engineering, Northwestern University, Evanston, IL; Department of Physical Medicine and Rehabilitation, Northwestern University, Chicago, IL; Shirley Ryan Ability Lab, Chicago, IL

## Abstract

Intracortical microstimulation (ICMS) of somatosensory cortex evokes tactile sensations whose location and properties can be systematically manipulated by varying the electrode and stimulation parameters^1–3^. This phenomenon can be used to convey feedback from a brain-controlled bionic hand about object interactions. However, ICMS currently provides an impoverished sense of touch, limiting dexterous object manipulation and conscious experience of neuroprosthetic systems. Leveraging our understanding of how these sensory features are encoded in S1^4,5^, we sought to expand the repertoire of ICMS-based artificial touch to provide information about the local geometry and motion of objects in individuals with paralysis. First, we simultaneously delivered ICMS through multiple, spatially patterned electrodes, adopting specific arrangements of aligned projected fields (PFs). Unprompted, the participants reported the sensation of an edge. Next, we created more complex PFs and found that participants could intuitively perceive arbitrary tactile shapes and skin indentation patterns. By delivering patterned ICMS sequentially through electrodes with spatially discontinuous PFs, we could even evoke sensations of motion across the skin, the direction and speed of which we were able to systematically manipulate. We conclude that appropriate spatiotemporal patterning of ICMS inspired by our understanding of tactile coding in S1 can evoke complex sensations. Our findings serve to push the boundaries of artificial touch, thereby enriching participants’ conscious sensory experience from simple artificial percepts to highly informative sensations that mimic natural touch.

## Main

Touch conveys rich information about our interactions with objects that facilitates dexterous behaviors^6^. When we touch and manipulate an object, we rapidly determine whether it is hard or soft, rough or smooth, sticky or slippery. Furthermore, we identify local contours or ridges under our fingertips, such as the edge of a button. This information is consolidated into a three-dimensional representation of the object in a phenomenon known as *stereognosis*^7^. This is enabled, in part, by neurons in primary somatosensory cortex (S1) that have receptive fields explicitly tuned for these features^8^. Spinal cord injury often disrupts both motor and sensory functions of the hand^9^. This loss can be partially circumvented by restoring hand and arm movements using robotics and decoding motor intent from motor cortex^10^, but dexterous hand function will be impossible without somatosensation. Intracortical microstimulation (ICMS) of Brodmann’s Area 1 (BA1) within the somatosensory cortex has been shown to evoke vivid tactile sensation on the skin^11–13^ and provides a promising avenue for restoring somatosensory feedback. ICMS sensation qualities, however, vary widely depending on the electrode, with reported sensations spanning from touch and pressure to tapping and tingling sensations^3,12^.

Initial ICMS attempts have been focused almost entirely on two features: sensation location^11–15^ and intensity^2,16^. ICMS can be used to intuitively signal the location of object contact on a bionic hand by linking sensors on the hand to somatotopically-matched electrodes in BA1^1^. The resulting sensations appear to originate from the corresponding skin location^11–14^. The perceived intensity - signaling interaction force - can be controlled by ICMS amplitude^11^ or frequency^3^. While contact location and force are critical feedback components, the sense of touch is far richer than this, also conveying information about the texture, material properties, local contours, and about the motion of objects across the skin. These aspects of touch are critical for our ability to flexibly and dexterously interact with objects (**Figure 1A**). Without these rich sensations, artificial touch will remain highly impoverished.

**Figure 1.**
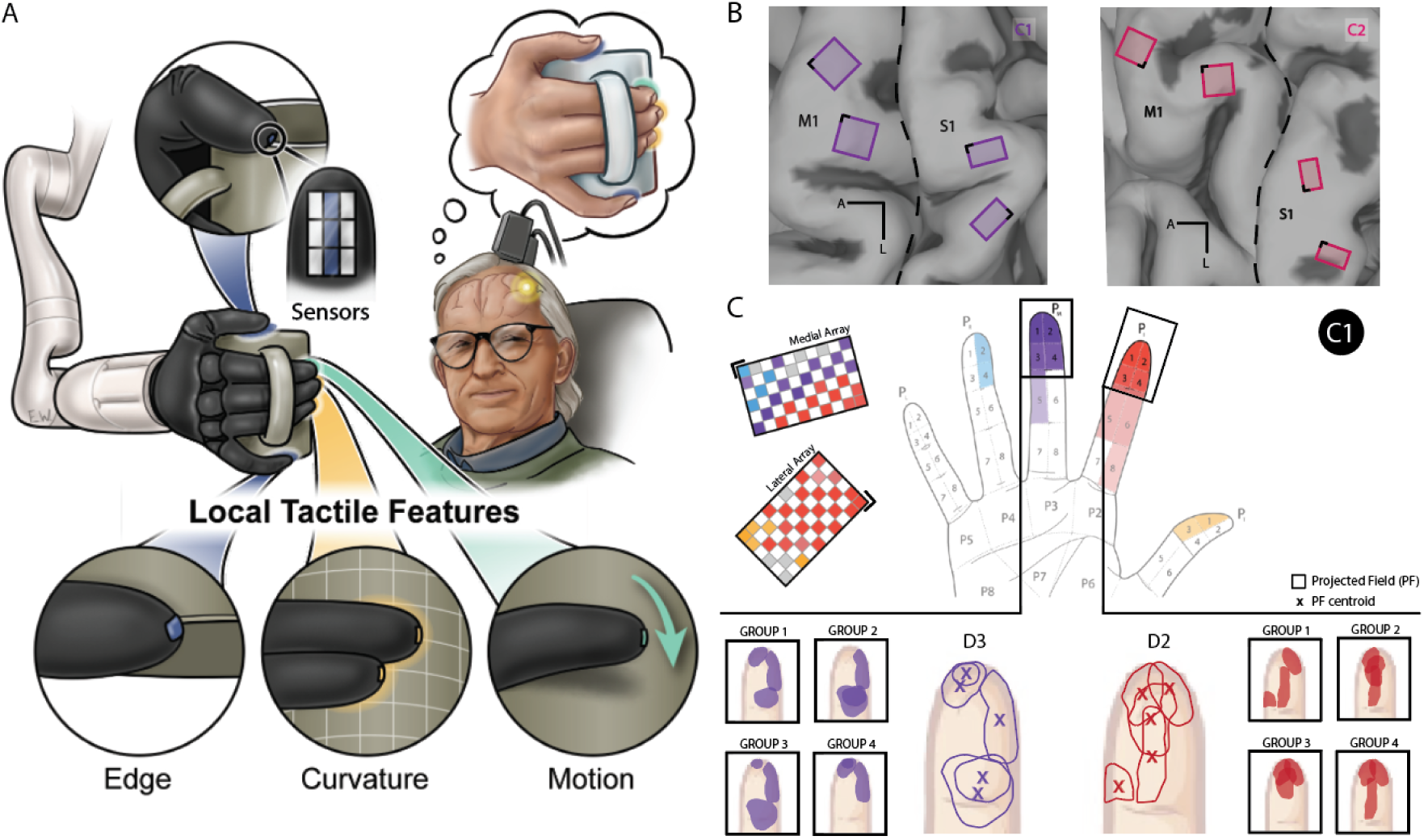
ICMS of S1 evokes tactile projected fields whose location follows the expected somatotopic organization. **A|** Diagram of ICMS-based somatosensory feedback concept. First, the sensorized robotic hand collects the tactile features from the local hand-object interactions. Second, spatio-temporal patterned ICMS of S1 is adopted to artificially encode these local features. Finally, sensory integration allows the user to recognize the grasped object through artificial touch. **B|** Four Utah arrays (Blackrock Neurotech, Inc.) were implanted in participants C1 and C2, two of which were placed in the hand representation of S1 (Brodmann’s area 1), based on localization with brain imaging techniques (fMRI and/or MEG). Here we show array locations based on intra-operative photos, superimposed on a pre-surgical anatomical MRI. The central sulcus is indicated by the dashed line. M1: motor cortex. S1: somatosensory cortex. L: Lateral. A: Anterior. **C|** Locations of projected fields – the location on the hand where sensations are experienced – for each S1 channel for participant C1. The top array is medial, bottom one lateral. Colors denote the location of the projected field. Gray squares denote electrodes that evoke sensations on the dorsum of the hand, and white squares denote unwired electrodes. Black corners indicate alignment. Zoomed parts show precise location of single electrode PFs on D2 and D3 and the possible combinations (groups) tracing different tactile shapes.

Leveraging the fact that simultaneous multi-electrode ICMS (mICMS) can evoke percepts that combine approximately additively across electrodes with overlapping PFs^14^, we sought to extend this phenomenon to PFs with specific configurations (e.g., alignment). Further, leveraging the fact that neural activity in BA1 evolves in a somatotopically predictable spatiotemporal way during object motion across the skin^8^, we asked whether structured spatiotemporal ICMS can recreate a sense of tactile motion. Together, we aimed to expand the repertoire of touch sensations beyond basic features to improve the sensory experience of bionic hands.

### Spatial patterning of ICMS evokes tactile shape percepts

We performed a series of experiments in two participants (C1, C2) with cervical spinal cord injuries, using microelectrode arrays implanted in the hand representation of BA1 (**Figure 1B**), the locations of which were based on pre-operative imaging. In both participants, ICMS resulted in distinct tactile sensations on the contralateral hand^11,14^ (**Figure 1C**). In these experiments, we identified combinations of three electrodes that had spatially aligned PFs which we hypothesized would evoke edge-like sensations (i.e., a composite tactile percept). Indeed, the participants, without knowledge of the intended goal of the experiment, spontaneously reported sensations with specific orientations and shapes on the skin (**Movie S1**). To test the robustness and utility of this phenomenon, we repeated this paradigm with PFs distributed along the length of a digit, across the digit, or with a random pattern (**Figure 2A**). The participants could reliably discriminate the orientation of the edge (average performance of 85% in C1 and 65% in C2, chance 33%, n=450 for C1 and n=150 for C2, Fisher Exact test, p<0.05). This result was consistent across digits (D) (C1: D1 – 81.3%, D2 – 89.3% and D3 – 84%, C2: D2 – 68% and D3 – 61%, chance 33%, n=150 for C1 and n=75 for C2, Fisher Exact test, p<0.05) (**Figure 2B**).

**Figure 2.**
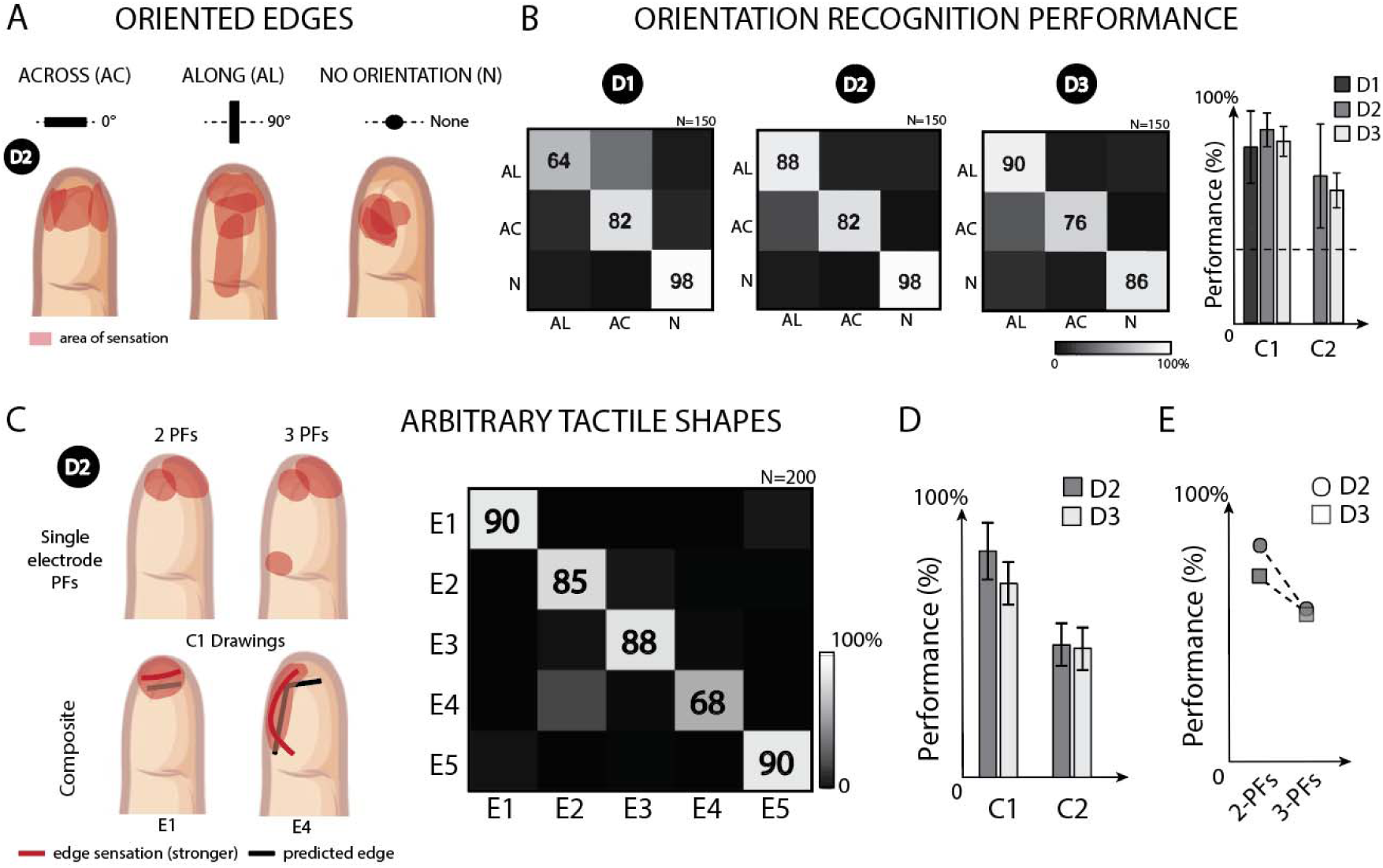
Multichannel ICMS of S1 evokes sensation of arbitrary tactile edges. **A|** Schematic of spatial organization of the stimulated electrodes evoking sensations on digit D2 (indicated in black circle) of Along (AL), Across (AC) and No orientation (N) edges on the microelectrode array and their corresponding projected fields. **B|** Orientation identification accuracy for each digit. Columns correspond to the true label of the stimulated set of electrodes, and rows to sensation identified by the participant. Numbers in the boxes indicate the percent of trials this stimulus was identified out of the times it was presented. N=150. Data from C1. On the right, overall orientation recognition performance for each digit in both participants. Error bars indicate confidence intervals. **C|** Examples of combinations of 2 single electrode PFs to create arbitrary tactile shapes on D2. Shaded area represents the area of sensation. Thick lines indicate the zone where the sensation is stronger/more intense. Black lines are the predictions of the evoked sensation from the single electrode PFs. Composite percepts reported by C1 when stimulated with the relative combination. Identification performance of five arbitrary tactile shapes, combining 2 PFs on D2, randomly presented to C1. Numbers in the boxes indicate the percent of success identifications per condition. **D**| Arbitrary shapes recognition broken down by participant (C1 and C2) and digit (D2, D3) (bottom left). Chance levels: C1 - 20% and C2 – 33%. **E|** Arbitrary shapes recognition broken down by ICMS-evoked edge complexity (2 and 3 combined PFs). Chance levels: 2PFs – 20% and 3PFs – 25%. (bottom right). Data from C1.

Next, we assessed the effect of ICMS duration and amplitude on the ability to determine edge orientation. Increased duration improved orientation discrimination, reaching asymptotic performance at 0.5 seconds (C1: 0.5 seconds – 78%, n=225, Fisher Exact test, p<0.05, **Extended Data Figure 1A**). The effect of duration on discrimination is consistent with the processing of natural touch^17^, particularly in absence of vision^18^. Moreover, it highlights the fact that feedback from bionic hands could signal local features in well under a second, making real-time feedback and control feasible. Unlike duration, there was no effect of amplitude on discriminability (C1: 40µA – 70%, 60µA – 67.7%, 80µA – 72.3%, chance 33%, n=270, Fisher Exact test, p>0.05, **Extended Data Figure 1B**), consistent with natural touch processing^19^. In other words, even if the intensity of the perceived sensation is modulated, its spatial properties remain recognizable and discriminable by the user.

We extended this paradigm to arbitrary angles and simple shapes, using a classification task in which participants matched mICMS sensation to shapes shown on a screen (3 and 5 shapes shown for C2 and C1, respectively). Both participants were able to correctly identify shapes both on D2 (C1: 82 ± 9.2%, chance 20%; C2: 50 ± 14%, chance 33%; Fisher Exact test, p<0.05) and D3 (C1: 72 ± 8%, chance 25%; C2: 48 ± 8%, chance 33%; Fisher Exact test, p<0.05) (**Figure 2C, D**). As expected, participants were more likely to confuse similar shapes (i.e., version of the same shape evoked through partially overlapping sets of electrodes) (**Extended Data Figure 2A**). Next, we tested more complex shapes composed of 3 PFs, often with two adjacent edges (**Extended Data Figure 2B**). Participant C1 was able to correctly recognize the 3-PFs shapes despite their greater complexity (60 ± 22.7% on D2, chance 25% and 57 ± 7.6% on D3, chance 25%, Fisher Exact test, p<0.05, **Figure 2E** and **Extended Data Figure 2C**). Performance was slightly reduced with tactile shape complexity as observed in natural touch^18^. In addition, in order to measure the degree to which tactile lines experienced by the participants matched those predicted by the single-electrode PFs, we had them draw the perceived shape (**Extended Data Figure 2A** and **Extended Data Figure 3**). We found that both the length and angle of the evoked tactile shape significantly correlated with our predictions (**Extended Data Figure 2D**-F).

In order to determine how the spacing between PFs influenced the percept of a continuous edge, we asked our participants to report if pairs of simultaneously stimulated electrodes were perceived as one continuous sensation or two discrete sensations. We found that multichannel PFs on the fingertip seem to be perceived as a composite continuous sensation, without spatial gaps, when separated by less than the distance of natural two-point discrimination thresholds (between 2 to 8 mm on fingertips^20^, **Extended Data Figure 2A**,B and **Extended Data Figure 3**). This is in line with theories based on the funneling illusion^21,22^ suggesting that cortical activation corresponds to the perceived, rather than actual, site of peripheral stimulation and that spatial perceptions are strongly dictated by central representations. Indeed, this stimulation of multiple electrodes could lead to spatial integration and a single cortical activation zone^22^.

The results presented so far involve simple flat/planar sensations, however, objects often have curves that indent the skin by varying amounts and mechanoreceptor activation depends on the specific curvature and the local shape^23^. Mechanoreceptors in the glabrous skin are sensitive to the edges of stimuli^24^, suggesting that the effective stimulus for these receptors is the curvature of the skin. The skin deformation (and the points of pressure) changes according to the level of curvature and the contact area. We explored our ability to convey the sensation of convex and concave surfaces by modulating stimulation amplitude and timing of electrodes with adjacent PFs. We delayed ICMS at the sites corresponding to the skin surfaces that would be contacted later, mimicking the temporal activation pattern of natural touch (**Figure 3A**). During the experiment, participant C1 reported sensations of rounded edges (**Movie S2**) and was able to reliably differentiate between concave, convex, and flat curves (64 ± 6.9%, chance 33%, n=75, Fisher Exact test, p<0.05, **Extended Data Figure 4**). To test the precision with which C1 could detect curvature, we chose electrodes with aligned PFs with varying curvature (from 0% to 50%, defined as relative difference in amplitude among the adopted electrodes). Task performance increased with curvature (both concave and convex); the participant could reliably identify (>75% success) curved stimuli once curvature exceeded 20% (**Figure 3B**), marginally worse than natural touch. In fact, the human tactile discrimination of curvature is around 10% both for concave and convex objects^25,26^ but depends on the contact areas between the curved surfaces and the finger pad skin^25^.

**Figure 3.**
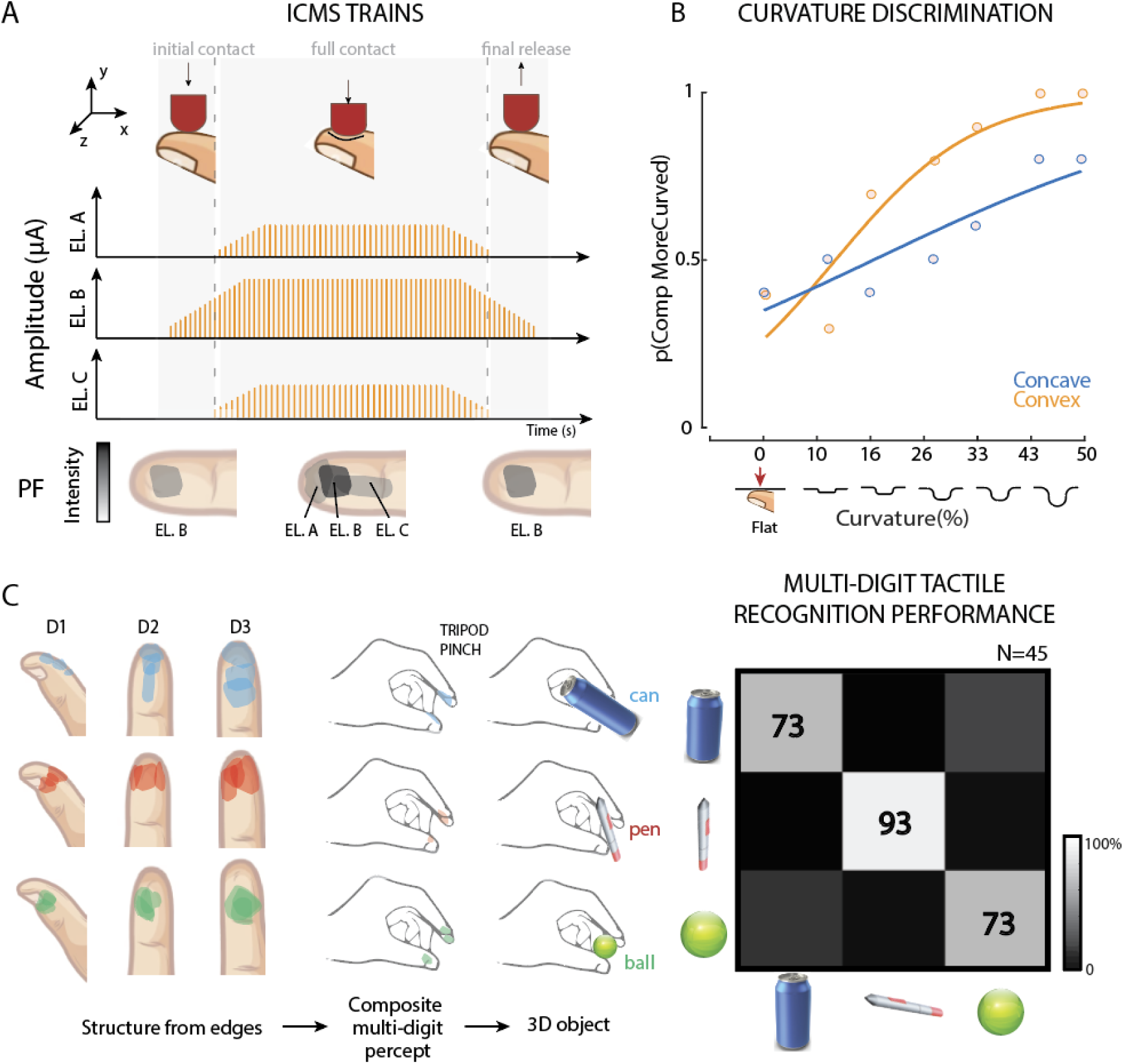
Tactile curvature and composite multi-digit tactile percepts via ICMS. **A|** Encoding edge curvature through spatiotemporal pattering of ICMS. Electrodes with aligned PFs are chosen to evoke a percept with a specific orientation. Then amplitude and synchrony between the activated channels are modulated in order to encode convex or concave stimuli on D2. **B|** In a curvature discrimination task, C1 was asked to compare the curvature of a flat surface to other with different curvatures (from 0% - flat to 50% - most curved). N=65. **C|** Combinations of PFs simultaneously activated on three different digits (D1, D2 and D3). PFs arise from single channel stimulation. Conditions represent stimuli on two perpendicular axes (Across and Along) and one condition with no orientation. mICMS evoked sensations on all three digits simultaneously. C1 reported to perceive to grasp real objects (Can, Pen and Ball). Multidigit identification performance. N=45. Numbers in the boxes indicate the percent of success identifications per condition. Data from C1.

Our initial experiments were restricted to PFs on a single digit, but when we grasp an object, multi-digit cutaneous inputs are integrated to form a representation of the object that is matched against memory and perception^27^.To determine if our approach could be extended to produce sensations of this complexity, we used mICMS that spanned PFs on multiple digits. Interestingly, C1 evoked sensations that he associated with grasping real objects (**Movie S3**). The combination of three edges across the fingers evoked sensations reminiscent of natural contacts: a pen when the three encoded edges on each finger were colinear, a can (grabbed from the top) when the edges were all along the length of the digits, and a ball, when the PFs evoked on multiple digits were all overlapped (no alignment) (**Figure 3C**). To test whether these evoked sensations were reliable, we designed a multichoice task in which we randomly presented 3D objects encoded on 9 channels with PFs on multiple digits. C1 successfully identified the encoded objects above chance (79 ± 11.5%, chance 33%, Fisher Exact test, p<0.05, n=45). Our findings demonstrated how informed patterns of ICMS could evoke vivid sensory experience associated with three-dimensional structures related to real-life objects.

### Spatiotemporal patterning of ICMS evokes sensations of tactile motion

Static spatial information, conveyed as described above, could improve object recognition for people with impaired somatosensation. However, stereognosis typically requires active manipulation to fully map the features on an object and build its internal representation. Consequently, we added a temporal component to our mICMS by sequentially activating channels with adjacent PFs. In both vision and touch, information about motion is extracted from a spatiotemporal pattern of activation across a two-dimensional sensory sheet (in the retina and skin, respectively), a process that has been extensively studied in both modalities^5^. In natural touch, perception of continuous motion of an object across the skin is enabled by smooth transition of activity between adjacent skin receptors^4,28^. The perceived motion direction is determined by the responses of a subpopulation of cortical neurons in BA1^8^. We asked if we could mimic that effect by stimulating electrodes with adjacent PFs in a specific temporal sequence. Indeed, electrodes are located at discrete locations on the cortex (BA1), requiring specific combinations to stimulate in a continuous trajectory. In both participants, we selected electrodes that formed two axes: proximal-distal – PD – (along the finger) and radio-ulnar – RU – (across the finger), which allowed four directions of motion (PD, DP, RU, UR). In C1, both axes shared a central electrode (PF), thereby forming a cross on the index finger pad (**Figure 4A**). In C2, the radio-ulnar movements were entirely on the middle finger, and the proximal-distal movements had PFs across index, middle, and ring finger (**Extended Data Figure 6A**). Each electrode was stimulated for 500 ms, with a 10 ms delay prior to the next electrode. Both participants immediately described the evoked sensation as “something moving on my fingertip” or “I am feeling like I am rolling my finger on a surface” (**Movie S4**), and both were able to distinguish the four directions of motion (C1: 76 ± 13.8 %, chance 25%, n=160, Fisher Exact test, p<0.05, C2: 78 ± 14.7%, chance 25%, n=60, Fisher Exact test, p<0.05, **Figure 4A** and **Extended Data Figure 6A**, top), including when stimuli spanned multiple digits (C1: 98 ± 2.5%, n=120; C2: 75 ± 14%, n=75, Fisher Exact test, p<0.05, **Figure 6A**, bottom and **Extended Data Figure 6B**). We were able to evoke other apparent motions with participant C1, including circular (i.e., clockwise, counterclockwise and rectilinear motion) and radial (contraction and expansion and static sensation) (Circular motion: 96 ± 7.5%, chance 33%, n=90, Fisher Exact test, p<0.05; radial motion: 81 ± 16.6%, chance 33%, n=75, Fisher Exact test, p<0.05, **Extended Data Figure 7A**,B).

**Figure 4.**
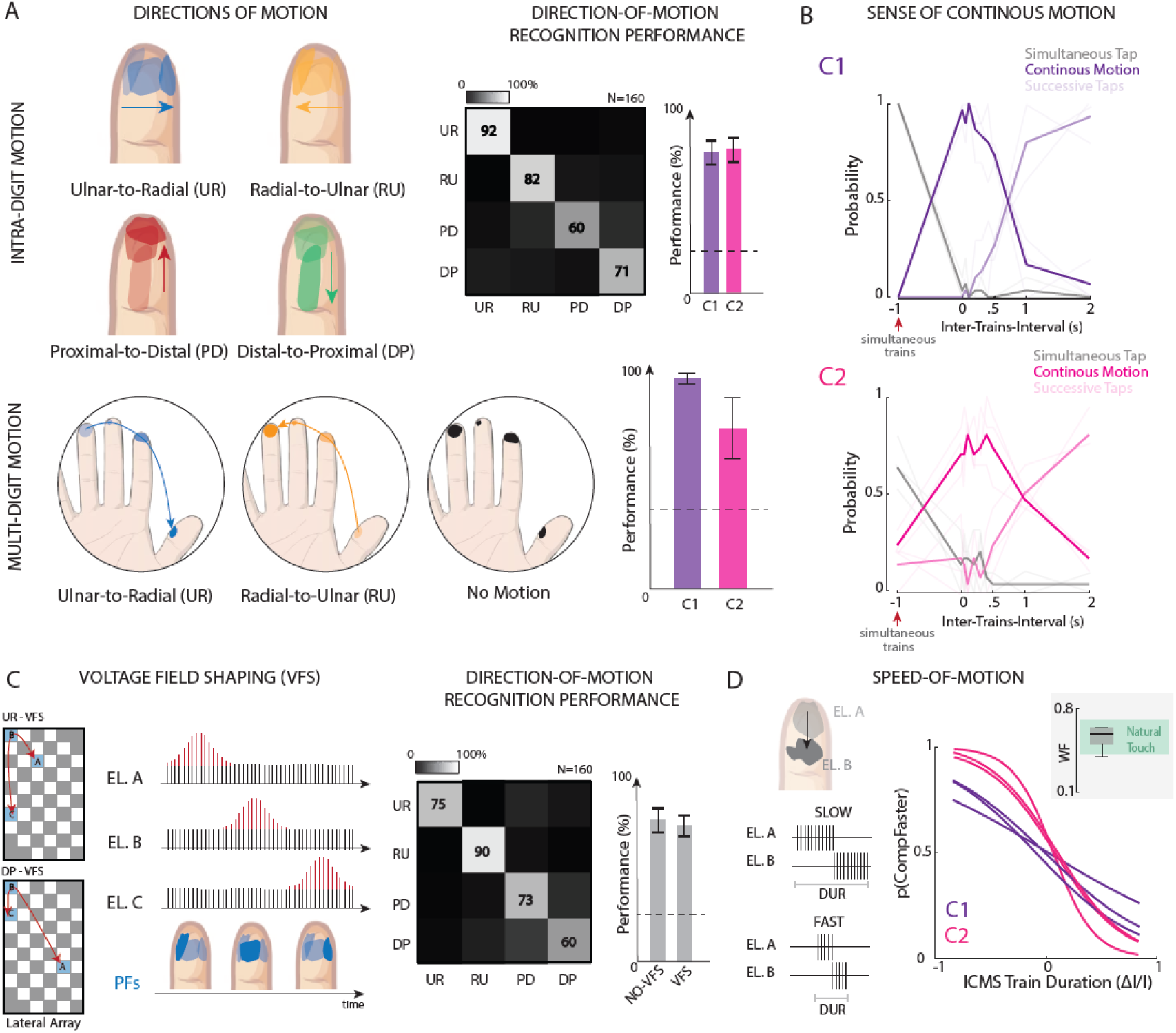
Patterned ICMS of S1 evokes sensations of apparent motion. **A|** (top) Combinations of PFs sequentially activated in four different directions (Ulnar-to-Radial, Radial-to-Ulnar, Proximal-to-Distal and Distal-to-Proximal). PFs relate to single channel stimulation. Direction-of-motion recognition performance. Numbers in the boxes indicate the percent of success identifications per condition. N=160. Performance broken down by participant (right). Chance levels: 25%. (bottom) Multi-digit motion recognition among 2 directions (UR-RU) and no motion in C1 (N=120) and C2 (N=75). PFs adopted in C1. Chance level: 33%. **B|** Perception of continuous motion. Two PFs in three configurations are simultaneously (inter-trains-interval=-1s) or sequentially (inter-trains-interval between 0s and 2s) activated. Thick Colored Lines represent sense of continuous motion (percent); Light Colored Lines represent sense of intermittent motion (taps); and Black Lines represent no motion (simultaneous tap). N=300. C1 and C2 data. **C|** Modulation of amplitude among multiple channels allows for shaping the applied voltage. Direction-of-motion recognition performance adopting amplitude modulation. Three PFs are involved in the encoded movements. Numbers in the boxes indicate the percent of success identifications per condition. N=160. Performance broken down by stimulation condition (NO-VFS and VSF) (right). Chance levels: 25%. Data from C1. **D|** Encoding motion speed through the modulation of the ICMS train duration between electrodes with different PFs. Speed discrimination performance for both C1 and C2 are reported. All the Weber fractions (WFs) are displayed in relationship with the natural range of our skin perception^37^ (green zone).

We assessed the effect of ICMS train duration and amplitude on the participants’ ability to determine direction of motion. While duration had a significant effect (50ms – 30%, 200ms – 72%, chance 25%, Fisher Exact test, p<0.05, **Extended Data Figure 5A**, with fixed inter-train-spacing), as with natural touch^29,30^, amplitude did not (40 - 60 – 80 µA, **Extended Data Figure 5B)**. As with edge orientation discrimination (**Extended Data Figure 1A**), only few hundreds of milliseconds were required to interpret motion direction, making this practical for a closed-loop BCI. Notably, apart from their relative locations on the skin, both edge orientation and motion encoding were achieved regardless of the specific characteristics of the individual PFs (quality, geometry, size, or perceived intensity).

To understand how the relative timing of ICMS trains might influence motion percepts, we systematically varied their degree of overlap (negative inter-train interval) or spacing (positive inter-train interval). We asked the participants to report whether a perception was continuous or pulsatile (**Figure 4B**). The perceived motion occurred only when trains were separated by 0 to 0.5 seconds. If trains were delivered with temporal overlap, they appeared to be a single event at two locations; if they were more than 0.5 seconds apart, the mICMS was typically perceived as two successive events (intermittent motion). This result was held regardless of the PF locations and spatial separation. These timing limits closely match those observed in natural touch^31^. Longer inter-train-intervals generated percepts described as two successive skin taps, rather than motion, consistent both with natural touch and vision (sequential taps^31^ or flashing lights^32^, respectively).

Searching for local landmarks and features is another important aspect of object exploration. When we grasp an object, neurons with the receptive field on the contact area start to fire, and then when that same object moves over the skin in a certain direction, our sensory system is able to encode that motion via temporal modulation of the firing^33,34^.This scenario refers to when, instead of sensing an object moving on the skin (motion only), we actively move our finger over a surface sensing local structures (contact + motion). To mimic this, we investigated the possibility of using more efficient dynamics of ICMS to minimize charge injection beyond fixed-amplitude trains like those proposed for intracortical visual prostheses (i.e., dynamic current steering – DCS – or voltage field shaping)^35^ and cochlear implants^36^. To encode this scenario, in C1 we simultaneously stimulated three electrodes with aligned PFs and we shaped the voltage fields across these three electrodes (VFS, **Figure 4C**). In particular, we modulated the amplitude of mICMS across electrodes having adjacent PFs, moving the amplitude peak in the direction of the desired apparent motion. With this spatiotemporal modulation of ICMS, the participant was able to identify motion among 4 directions on the skin (D2) (C1: 74 ± 12.3%, n=160, Fisher Exact test, p<0.05, **Movie S5**). Discrimination performance was not statistically different compared to sequential mCMS (NO-VFS, Fisher Exact test, p>0.1). This result demonstrates that this encoding method enables the simultaneous disentanglement of three distinct tactile features from ICMS: location, intensity, and direction of motion encoded in the artificial percept.

Speed variation is another fundamental feature of tactile motion, crucial for encoding object interactions such as slippage or surface exploration^6^. To explore this feature, we varied the temporal activation of two adjacent PFs, and asked the participants to identify which of two mICMS patterns they perceived to be faster (**Figure 4D** and **Movie S6**). Both participants were able to discriminate motion speed with a Weber Fraction of 0.56 ± 0.09 for both C1 and C2, which falls in the same range as natural touch^37^. In other words, the speed sensitivity of artificial stimuli is similar to that of tactile stimuli applied directly on the skin. In addition, ICMS trains with different stimulus-onset lags (i.e., the time between the start of two sequential stimulus trains) were perceived as different speeds both on D2 and D3 in C1 (Weber fraction: 0.63 ± 0.06, **Extended Data Figure 8B**). To ensure that the participants were identifying speed of motion rather than train duration, we repeated the speed discrimination task using electrode pairs with different distances between PFs, while keeping the ICMS train durations unchanged (**Extended Data Figure 8A**). We selected channels with short (∼0.6cm) and long (∼1.4cm) inter-centroid distance on D2 and interleaved both as standard stimuli. We intermixed them in the same experimental block. When both the standard (500ms) and comparison (79 - 921ms) stimuli were presented at the same distance (both short and long), there was no significant difference in PSE (i.e., point-of-subjective equality). In contrast, ΔPSE was 360ms (PSE speed: 1.63cm/s) and -140ms (PSE speed: 1.67cm/s) for short and long PF separation, respectively, confirming that the effect is one of perceived speed, not train duration (Δdistance / ICMS_duration - cm/s). In the context of natural touch, the neural representation of motion speed could be based on a within-fiber intensity code^38^ and on a between-fiber spatiotemporal code^39^. The latter is defined as the speed of the sequential activation of neighboring afferent fibers in response to a traceable feature of the skin surface, moving across their receptive field^39^. The developed ICMS encoding scheme resembled the afferent spatiotemporal code of motion on the skin surface.

### Tracing trajectories on the cortex allows complex tactile shapes to be encoded

Motion is also an important component of tactile discrimination. Complex tactile patterns, as letters, are easier to recognize when they are drawn on the skin rather than simply indented^40^ (i.e., graphesthesia), and active exploration of objects is more informative than static deformation of the skin^41^. Similarly, shapes traced through the visual field using sequential mICMS within V1 are better recognized than are the corresponding simultaneous ICMS stimuli^35^. To test the limits of our ability to convey complex tactile shapes and the effect of motion on tactile discrimination, we compared PFs representing letters that were activated either *sequentially* or *simultaneously*. Participant C1 was able to identify the letters T, L, C, O, I represented on 3 - 6 adjacent PFs on D2(**Figure 5A**) with both simultaneous and sequential mICMS. However, performance was significantly higher when the electrodes were activated sequentially (48 ± 18.6% compared to 37 ± 21.3%, n=150, chance 20%, Fisher Exact test, p<0.05, **Figure 5B**). The advantage of sequential activation increased with increasing shape complexity (number of bars composing the letter; **Figure 5C**). While we tested only letter-like shapes, the outlines of other common objects could also be traced using the same principles.

**Figure 5.**
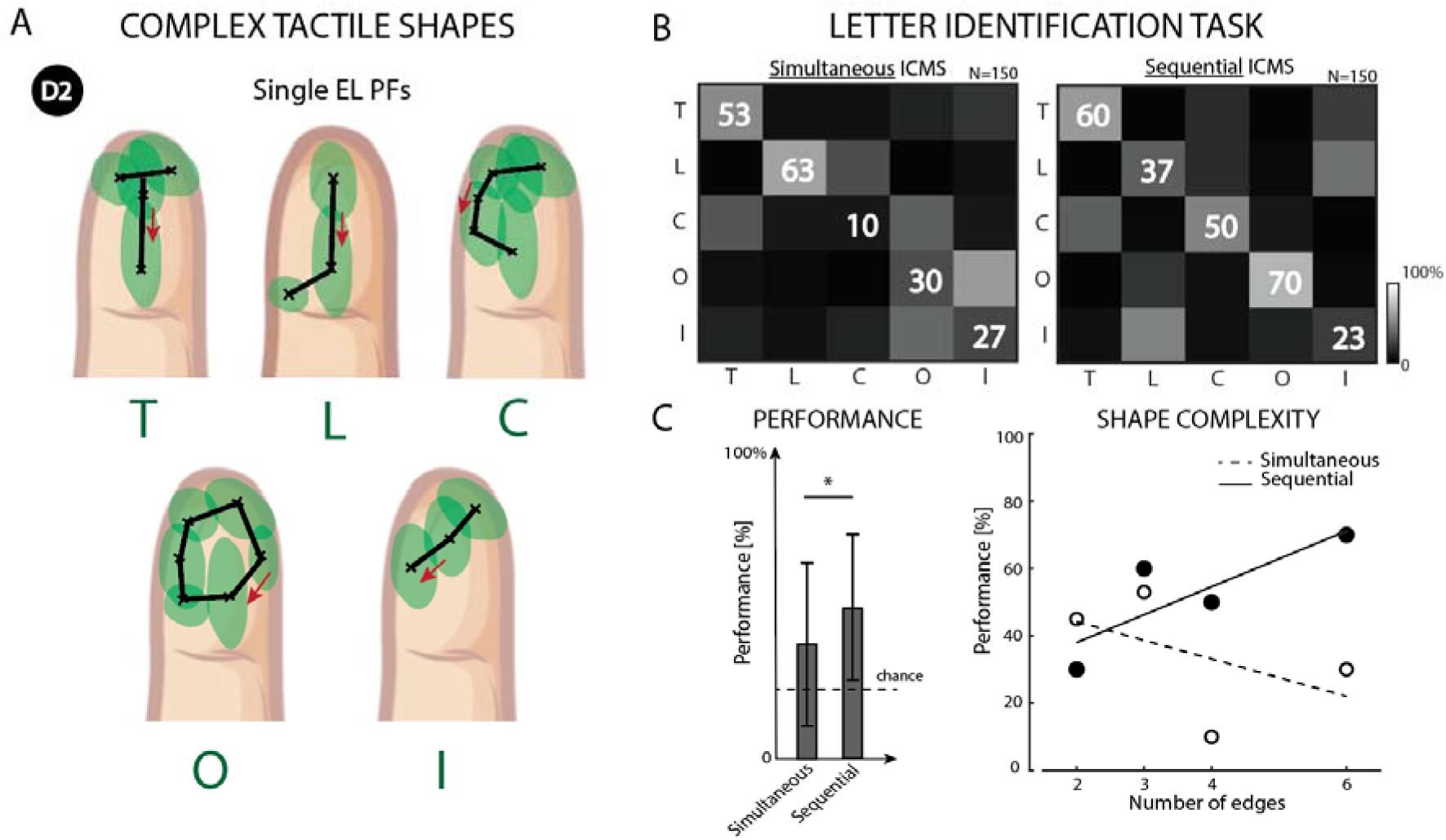
Sequential versus simultaneous multichannel ICMS for encoding complex tactile shapes. **A|** Selection of PFs (>3) creating letters on D2. x indicate single electrode PF centroids. Red arrows show the order of sequential electrode activation. **B|** Letter recognition task encoded using sequential or simultaneous ICMS. Numbers in the boxes indicate the percent of success identifications per condition. N=150. Data from C1. **C|** Identification performance according to shape complexity (right) and direct comparison between sequential and simultaneous ICMS (left). *p<0.05.

Sequential activation also has the benefit that it reduces the current injected into the cortical tissue at any given point, decreasing sensory adaptation and reducing the likelihood of epileptic seizures due to synchronized activation of multiple electrodes^35^. When the complexity of the tactile shape increases, the resulting PFs can interact, often coalescing into a single PF that is not always described as a specific shape (but more as a general area of sensation) (**Figure 5C**). Spatiotemporal mICMS seems to diminish the likelihood of spatial blurring, improving the effective spatial resolution of the patterns that can be evoked. In addition, this encoding strategy might evoke a pattern of neural activation more similar to the processing of natural tactile motion. Information about direction of motion and speed are transmitted also thanks to local transients of neural firing modulated across neurons with adjacent receptive field^8^, similarly as this modulation of mICMS amplitude. In the field of somatosensory neural prosthetics, the use of more biomimetic approaches to stimulate the nervous system showed promising effective benefits compared to canonical stimulating strategies^2,42,43^.

## Discussion

This study represents the continuation of decades of experiments exploring stimulation of both human and non-human primate somatosensory and visual cortices toward the goal of building cortical neuroprostheses. Despite the presence of neurons in the visual cortex tuned to specific features (e.g., orientations^44^), electrical stimulation causes only simple, typically white, phosphenes (the visual equivalent to a tactile PF), the result of activating all neurons in the proximity of the electrode^45^ regardless of their feature selectivity^46^. However, despite this limitation, mICMS applied to primary visual cortex has been used to evoke coherent visual forms^47–49^. In the somatosensory cortex as well, single electrode stimulation evokes a direct activation of all the surrounding neurons characterized by a short latency excitatory response followed by a period of inhibition, regardless of the specific stimulus selectivity^50^. Neurons in BA1 have inputs organized to select specific stimulus features, such as (a) contact area, (b) edge orientation, (c) motion across the skin, or (d) direction of movement. These feature detection neurons have larger receptive fields than those of Area 3b and require inputs to be presented in specific spatial and/or temporal configurations^36^. More complex spatiotemporal mICMS may be an effective means of activating higher-order percepts that more closely mimic the activation of somatosensory cortex. Our findings show that the perceived edge length correlated significantly with the separation (i.e., cortical distance) between electrodes (**Extended Data Figure 9A**). But beyond this, the arrays’ placement adopted in this study, based on cortical somatotopy^51^, also made it possible to configure specific PFs; edge length and orientation are functions not only of the number of stimulated electrodes, but also of their spatial distribution.

The sense of touch also allows perception of three-dimensional shapes. In the absence of vision, tactile object perception includes the integration of specific tactile information including contact points, curvatures, texture and local edges^26^. During stereognosis, both the cutaneous sense and the kinesthetic sense (input from receptors located in muscles, tendons, and joints) convey useful integrated information^52^. Previous studies have attempted to restore a sense of object shape, compliance or size using electrical stimulation of nerves^53,54^ and cortex^2,55^. However, three-dimensional shape encoding that leverages tactile features on multiple digits has not been previously explored. It should be noted that C1 received no proprioceptive feedback and could therefore imagine the hand aperture to be different during different trials, e.g. wide for a can and narrow for a pen. We hypothesize that when coupled with visual feedback, these artificial sensations will result in even more natural percepts due to the additional context.

Beyond these spatial components, when a sequence of natural stimuli contacts different points on the skin, they can cause activity patterns to cross the cortex, leading to apparent motion. The direction of motion reported by participants is the result of similar trajectories of mICMS-induced activity across the cortex (**Extended Data Figure 10A**-C). In the visual domain, as in our experiments, current delivered to electrodes in a sequence causes apparent motion^35^. Subpopulations of neurons in BA1 integrate local motion signals emanating from object contours and terminators to achieve a percept of global motion on the skin based on a vector average mechanism^8^, which can account for our ability to discern the direction of tactile motion. With spatiotemporal mICMS informed by these configurations, we evoked conscious and informative sensations of motion, including its direction and speed.

mICMS has been previously considered to be a promising approach for improving other properties of artificial touch. A reduction in detection thresholds and reaction times have been reported in both humans^15^ and monkeys^56^. Improved tactile discriminability and localizability has also been documented with the use of mICMS^2,14^. Moreover, mICMS can be used to provide a wider dynamic range of sensations, which can, for example, convey more levels of force^2^. However, a potential drawback of mICMS is its effect on motor decoding; S1 ICMS directly generates M1 activity^57^. This activity, not present in the decoder training data, causes decoding errors. To the extent that mICMS evokes even more activity in M1 than does single-electrode ICMS^57^ it will likely exacerbate the problem. This disruption, however, can be minimized by implementing biomimetic stimulation trains – which emphasize contact transients (through phasic ICMS) over maintained contact^1,58^. The relatively simple biomimetic spatiotemporal patterning of the mICMS used in these experiments could be expanded to convey even more complex tactile experience such as textures. In BA1, texture information carried in patterns of afferent activation are converted into a rate code by the spatial and temporal filtering properties of subpopulations of cortical neurons. These act individually as encoders of spatial and temporal features in the input^59^. Implementing these specific spatiotemporal patterns in the mICMS trains would eventually allow to encode complex sensations related to textures.

Hardware improvements are possible as well. Our S1 arrays consisted of 6x10 electrodes (with every other electrode wired in a checkerboard pattern) covering 2.4x4.0 mm, allowing only limited coverage of the hand. Many more electrodes will be required to cover the whole hand area; closer spacing would allow for more detailed maps of PFs. While implanting larger arrays would improve coverage, implanting more arrays improves the chances that all digits are represented while still avoiding penetrating blood vessels. Developing higher density arrays would allow activation of small volumes in a larger cortical area, maintaining the likelihood of interactions between adjacent electrodes low. These high-resolution maps are necessary to restore acuity and richness closer to natural touch. Furthermore, given that digit tip representations in area 3b are deep within the central sulcus, the only access to digit tip representations with the 1.5 mm long electrodes currently approved for human use is at the border of areas 1 and 2.

We have shown previously that it is possible to map the sensors on bionic hands to somatotopically-matched electrodes in S1 to yield intuitive feedback about the locations of object contact, with normal force used to dictate ICMS intensity^2,16^. To expand the repertoire of artificial touch and provide mICMS related to local edges and motions, bionic hands would require better sensorization, including smaller sensors and the ability to detect motion or tangential forces. For example, signaling an oriented edge requires a sensory sheet with sufficient resolution to detect its presence and orientation (**Figure 1A**). An expanded sensory capability will spur an increase in the density of the stimulating electrodes implanted in S1 such that the breadth of sensory features can be conveyed. The required spatial resolution of the sensors will be determined by the spatial resolution of the neural interface (the density with which available PFs will tile the hand). The development of highly sensorized artificial skin^60,61^ will help to exploit the full potential of ICMS and restore a more natural touch experience to the BCI users.

Summarizing, in this study, we designed, implemented, and tested novel neurostimulation paradigms which were able to combine artificial tactile percepts in a composite tactile experience. We tested them in individuals with spinal cord injury implanted with Utah arrays in the hand representation of their S1. We *simultaneously* or *sequentially* stimulated multiple electrodes to evoke complex tactile patterns. We demonstrated that the simultaneous stimulation of multiple electrodes in human S1 gives rise to the perception of shape and that dynamic stimulation gives rise to the perception of tactile motion. In the context of BCI, we showed that all evoked composite sensations occur on a time scale that are relevant for neuroprosthetic devices and also similar to natural touch. Therefore, we show that BA1 represents an effective implant location for restoring meaningful tactile experience. Considering the multidimensionality of tactile experience and the limitations of the current neural interfaces, this result was deemed not possible; perhaps requiring more complex technologies than currently exists, or perhaps totally impossible. Instead, we show here that by using principled neuroscientific understanding of the neural encoding of conscious sensory experiences, we could produce complex sensations with existing technology and feasible stimulation protocols. In the process, we have also suggested electrode and sensorization changes that might further improve the tactile perceptual experience of mICMS. Informed spatiotemporal ICMS has the potential to restore functional, life-enhancing touch in people with paralysis.

## Data Availability Statement

All data will be stored at the Data Archive BRAIN Initiative (https://dabi.loni.usc.edu/dsi/UH3NS107714) after publication and code for analysis is available upon request.

**Supplementary information** is available alongside this paper.

## Author contributions

Conceptualization: G.V., C.M.G., R.A.G. and S.J.B.; Development: G.V., A.H.A, R.L., L.J.E., D.P., and C.M.G.; Investigation: G.V., C.M.G., J.E.D., A.R.S., M.L.B., L.E.M., R.A.G., J.L.C., and N.G.H.; Analysis: G.V., C.M.G., and S.J.B.; Surgical implantation and Clinical Oversight: P.C.W.; Supervision: R.A.G. and S.J.B.; Writing: G.V.; Review and Editing: All Authors.

## Acknowledgments

We would like to thank the participants for their generous contribution to the advancement of science. We thank Prof. Sliman Bensmaia (who unexpectedly passed away in August 2024) for his incredible support, inspiring brainstorms, and unique passion during the study. Prof. Bensmaia rigorously investigated these features of natural touch in both humans and monkeys during his career. He inspired and pushed us to leverage what we learned from touch encoding to shape new ICMS paradigms in the context of human BCI. Research reported in this publication was supported by the National Institute of Neurological Disorders and Stroke of the National Institutes of Health under Award Number UH3NS107714, by R35 NS122333 and by the University of Chicago (Chicago Postdoctoral Fellowship). The content is solely the responsibility of the authors and does not necessarily represent the official views of the National Institutes of Health.

## Methods

### Participants

This study was conducted under an Investigational Device Exemption from the U.S. Food and Drug Administration and approved under a single Institutional Review Board protocol at the University of Pittsburgh. The clinical trial is registered at ClinicalTrials.gov (NCT01894802). Informed consent was obtained before any study procedures were conducted. Participant C1 (m), 55-60 years old at the time of implant, presented with a C4- level ASIA D spinal cord injury (SCI) that occurred 35 years prior to implant. Participant C2 (m), 60-65 years old at the time of implant, presented with C4-level ASIA D spinal cord injury (SCI) and right brachial plexus injury that occurred 4 years prior to implant. Thanks to the highly rich PFs maps of C1 having multiple aligned PFs on individual digits (**Figure 1B**), we tested experiments with more complex tactile shapes, curvatures and control tests than in C2.

### Cortical implants

We implanted four microelectrode arrays (Blackrock Neurotech, Salt Lake City, UT, USA) in each participant. The two arrays (one medial and one lateral array) in Brodmann’s area 1 of somatosensory cortex were 2.4 mm x 4 mm, with sixty 1.5-mm long electrode shanks wired in a checkerboard pattern such that ICMS could be delivered through 32 electrodes. The two arrays in motor cortex were 4 mm x 4 mm, with one-hundred 1.5-mm long electrode shanks wired such that 96 electrodes could measure neural activity. The inactive shanks were located at the corners of these arrays. Two percutaneous connectors, each connected to one sensory array and one motor array, were fixed to the participant’s skull. We targeted array placement based on functional neuroimaging (fMRI or MEG) of the participants attempting to make movements of the hand and arm, within the constraints of anatomical features such as blood vessels and cortical topography.

### Intracortical microstimulation (ICMS)

Stimulation was delivered via a CereStim 96 (Blackrock Neurotech). Each stimulating pulse consisted of a 200-µs cathodic phase followed by a half-amplitude 400-µs anodic phase (to maintain charge balance), the two phases were separated by 100 µs interphase.

### Simultaneous and Sequential Multi-channel ICMS

We selected groups of 2-7 electrodes. In most cases, the electrodes had aligned projected fields (the patch of skin over which the ICMS-evoked sensation is experienced). For C1, each of the selected electrodes evoked a PF in a single digit (D1, D2 or D3 with detection thresholds: 28.6 ± 15.6 µA). For C2, the selected electrodes were organized in groups (from 2 to 5) evoking then a PF in a single (D3) or multiple digits (D2-D3 or D3-D4). When stimulating through multiple electrodes, all electrodes delivered the same ICMS pulse train simultaneously for encoding edges. When attempting to evoke motion sensations, however, stimuli were delivered through electrodes sequentially. During each experimental block, we randomly interleaved stimulation through each electrodes’ combination.

### Projected fields – PFs

Projected fields were documented over multiple years for C1 or months for C2 for each electrode or electrode group. On each trial, a 60 µA, 100-Hz ICMS train was delivered through a given electrode or group and the participant drew the spatial extent of the sensation on a digital representation of participant hand. The participant could request as many repetitions of the stimulus as desired. The region enclosed by the drawn boundary constituted an estimate of the projected field for that electrode on that session. PFs obtained for each electrode or group were combined across sessions to obtain a time-averaged estimate (such as that shown in **Figure 1C**). From these digital images, we counted and also computed the PF centroid (center of mass). We then computed an aggregate PF for each stimulating channel by weighting each pixel on the hand by the proportion of times it was included in the reported PF over the duration of the study. This allowed us to estimate which combinations of electrodes would reliably evoke aligned PFs. See ^2,14^for more information.

### Edge Orientation Task

The edge orientation task was designed to assess participants’ ability to identify the orientation of tactile sensations induced by simultaneous mICMS. In this task, participants were presented with tactile sensations through multiple electrodes, each aligned along different axes (plus a random pattern). They were then asked to report the orientation of these sensations. Participants performed this discrimination task in an N-forced choice discrimination paradigm. On each trial, a stimulus lasting 2 sec was presented and the participant reported stimulus orientation. The order of presentation of the stimuli was randomized and counterbalanced. Data was obtained from each electrode combination over a minimum of 5 experimental blocks, each consisting of 10 presentations of each stimulus. The frequency of the ICMS stimuli was 100 Hz. This task was performed by C1 on D1, D2 and D3 and C2 on D2, D3. The accuracy of their responses was evaluated by comparing them with the actual orientation of the ICMS-induced percepts. The task was repeated in C1 varying both amplitudes (40, 60, 80 µA) and duration (250ms, 500ms, 1s, 2s, 3s) for each stimulus orientation. The stimulation parameters were the same on each electrode.

### Multi*-*channel PFs drawings

The multi-channel projected fields (PFs) drawing task aimed to explore how participants perceive and represent tactile percepts induced by mICMS. After receiving the desired number of repetitions of each stimulus, participants were asked to draw the perceived patterns on a digital hand representation. These drawings were then analyzed to assess their correlation with the expected tactile percepts, based on the stimulated electrode positions. This task provided insights into how mICMS can be used to convey complex tactile information. Single electrodes were chosen to evoke PFs on individual digit having the lowest detection thresholds for that specific patch of skin. The estimated shapes from mICMS are calculated based on the centroid locations of the correspondent individual PFs. A line passing through all centroids represented the estimated tactile shape. At the beginning of each session, each PF was double checked to confirm its local characteristics. Tactile shape complexity was varied from low complexity (hotspots from a single PF) to intermediate complexity (lines and curves from 2 or 3PFs), to “highly complex” percepts (letters from three or more PFs). To compare the ICMS-evoked edges with those estimated from the adopted electrodes, their predicted and perceived angles and lengths were calculated (Pearson’s correlation). Both C1 and C2 performed this experiment.

### Arbitrary shapes classification

The arbitrary shapes classification task, building off of the multi-channel PFs drawing task, investigates participants’ ability to classify various shapes encoded through combinations of different electrodes in mICMS. Different shapes were represented by specific electrode combinations, and after experiencing the induced sensations, participants were asked to classify these shapes from a list of visually displayed options. The accuracy of their classification was measured, providing valuable data on the effectiveness of shape representation through ICMS. The tasks were performed both on D2 and D3 in C1 and C2. Participants performed this discrimination task in an N-forced choice discrimination paradigm. On each trial, a stimulus lasting 2 sec was presented and the participant reported stimulus shape. The order of presentation of the stimuli was randomized and counterbalanced. Data were obtained from each electrode combination over a minimum of 4 experimental blocks, each consisting of 10 presentations of each stimulus. The frequency of the ICMS stimuli was 100 Hz and the amplitude 80 µA. The stimulation parameters were the same on each electrode.

### Curvature discrimination and classification tasks

For encoding tactile curvature, trapezoidal ICMS traces (2-sec in duration, 0.2-sec ramps) were delivered with a time offset on 3 electrodes having aligned PFs. On the electrode evoking PFs in between the other two electrodes, the ICMS trains started earlier and lasted more (0.2-sec). Moreover, its peak amplitude was also set higher (depending by the curvature - percentage). This was implemented to emulate the interaction of the finger pad with a curved object. Indeed, the receptive field activations would not be simultaneous, and the force applied on the skin would be centered on the curve peak. C1 performed a curvature discrimination task in a two-alternative forced choice paradigm. On each trial, a pair of stimuli, each lasting 2 sec, was presented with a 2-sec inter-stimulus interval and the participant reported which stimulus was more curved. The standard stimulus, consistent across the experimental block, was paired with a comparison stimulus whose curvature (amplitude and time occurrence of the central channel) varied from trial to trial and spanned the standard curvature (0% to 50%). The order of presentation of the standard and comparison stimuli was randomized and counterbalanced. Data were obtained over a minimum of 8 experimental blocks, each consisting of 2 presentations (one for each order) of each stimulus pair. The frequency of the ICMS stimuli was 100 Hz. To test whether ICMS curvature discrimination was subject to concavity, we selected convex and concave stimuli. The former was encoded with the central channel being activated earlier and set to higher amplitude while the latter adopted the opposite configuration. Curvature percentage represents the difference in amplitude and time onset between the selected channels. Tests for both concavities were interleaved and randomized within an experimental block. Then, for curvature recognition, C1 performed this task in a 3- forced choice discrimination paradigm. On each trial, a stimulus lasting 2 sec was presented and the participant reported stimulus concavity (Flat, Convex and Concave). The range of amplitude modulation for each combination was tuned per individual channels in order to evoke a matched perceived intensity. The order of presentation of the stimuli was randomized and counterbalanced. Data were obtained over a minimum of 5 experimental blocks, each consisting of 5 presentations of each stimulus. The frequency of the ICMS stimuli was 100 Hz.

### Object discrimination task

Multi-digit stimulation was provided to C1 combining the oriented edges presented in the Edge Orientation Task. Combinations of 9 electrodes (3 per digit) were adopted to encode different object shapes. Edges all with across orientations on three digits encoded a pen, along the fingertips’ orientations a can, otherwise a ball (random and overlapped patterns). For 3D object recognition, C1 performed this task in a 3-forced choice discrimination paradigm. On each trial, a stimulus lasting 2 sec was presented and the participant reported stimulus type (Can, Pen and Ball). The order of presentation of the stimuli was randomized and counterbalanced. Data were obtained over a minimum of 3 experimental blocks, each consisting of 5 presentations of each stimulus. The frequency of the ICMS stimuli was 100 Hz and the amplitude to 60µA on each channel.

### Direction of motion tasks

The Direction of Motion Tasks evaluates participants’ perception of the direction of motion from tactile percepts induced by sequential multi-channel ICMS. A single sequence of ICMS pulses across multiple (3) electrodes occurring in one of four directions (Ulnar-to-Radial, Radial-to-Ulnar, Proximal-to-Distal and Distal-to-Proximal) were used to simulate motion, and participants were tasked with reporting the perceived direction. Trains of 500ms at 60µA were sequentially delivered through aligned electrodes according to the specific direction. Both C1 and C2 performed this task in a forced choice discrimination paradigm. On each trial, a stimulus lasting 1.5 sec was presented and the participant reported stimulus direction. The order of presentation of the stimuli was randomized and counterbalanced. Data were obtained over a minimum of 3 experimental blocks, each consisting of 5 presentations of each stimulus. The frequency of the ICMS stimuli was 100 Hz. The task was repeated varying both amplitudes (40, 60, 80 µA) and duration (50ms, 200ms, 400ms, 600ms, 800ms) for each stimulus direction. The stimulation parameters were the same on each electrode. The same behavioral task was adopted for multidigit motion recognition, circular motion, and radial motion. In the multidigit task, participants were asked to identify the direction of motion across fingers. The same parameters were adopted for this task, while the adopted electrodes had PFs on different digits (D1, D2, D3, D4 for C1 and D2, D3, D4 for C2). In the radial motion encoding, the adopted channels evoked PFs oriented on a circular trajectory on D2 and then sequentially activated in clockwise, counter-clockwise or rectilinear (no rotation) order. For radial motion, the stimulation was provided sequentially from one channel and then from three channels for encoding expansion, vice versa for contraction and simultaneous stimulation on three channels was provided for static sensation (no radial motion). Circular and radial trajectories of motions were tested only in C1.

### Sense of continuous motion

The sense of continuous motion was assessed using a multichoice paradigm in both C1 and C2. Participants were asked to judge whether a stimulus was encoding a simultaneous tap, a continuous motion or two successive taps on the skin. To test this hypothesis, two PFs have been chosen and activated with different Inter-Trains- Intervals (ITIs). Since each train lasted 1s, -1s represent simultaneous delivering of the ICMS trains. ITIs of 0s, 0.05s, 0.1s, 0.2s, 0.3s, 0.4s, 0.5s, 1s, and 2s were tested. The order of presentation of the stimuli was randomized and counterbalanced. Data were obtained over a minimum of 1 experimental block, each consisting of 10 presentations of each stimulus. The frequency of the ICMS stimuli was 100 Hz and amplitude at 60µA.

### Spatiotemporal ICMS for motion encoding

Similarly to dynamic current steering (DCS or voltage field shaping), we used spatiotemporal ICMS to create virtual electrodes in between the physical electrodes on the array by using current steering. Voltage field shaping consisted of simultaneous stimulation of two electrodes in the sequence at a particular current ratio. The ratio is adjusted to change the location of the virtual electrode. The amplitude ratio varied for each individual pulse within the pulse train following a normal probability density function. The rate of change of the current ratio determines how rapidly the pattern is ‘‘drawn’’ on the cortex and how dynamic the pattern is perceived to be. In detail, three channels having aligned PFs and detection threshold below 40 µA were chosen for testing spatiotemporal ICMS. All three electrodes were activated using ICMS trains having a constant baseline at 40 µA for the entire length of the stimulus (1.5s). In addition to this stimulation, the amplitude was sequentially modulated on each channel following a normal probability density function with σ=0 for 500ms. In detail, each ICMS trains was designed to follow an amplitude envelope as a normal probability density function:

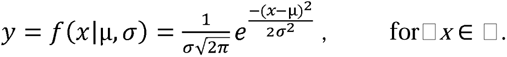

In this way, the voltage filed was shaped towards the next channel evoking a perception of apparent motion. The Direction of Motion Task was repeated using spatiotemporal ICMS. Data were obtained over 5 experimental blocks, each consisting of 8 presentations of each stimulus. The frequency of the ICMS stimuli was 100 Hz and amplitude modulated between 40 and 90µA following a normal probability density function. Data was collected in C1 using six PFs aligned along and across D2.

### Speed discrimination task

The Speed Discrimination Task tests participants’ ability to discern the speed of motion from different sequences of ICMS pulses. Two sequences with varying pulse durations are delivered across two specific electrodes (or groups of electrodes), simulating different movement speeds. Both C1 and C2 are asked to discern which sequence represents a faster motion on skin. Participants performed a speed discrimination task in a two-alternative forced choice paradigm. On each trial, a pair of stimuli, each lasting 1 sec, was presented with a 1-sec inter-stimulus interval and the participant reported which stimulus was faster. The standard stimulus, consistent across the experimental block, was paired with a comparison stimulus whose speed (ICMS train duration or stimulus onset) varied from trial to trial and spanned the standard speed. The order of presentation of the standard and comparison stimuli was randomized and counterbalanced. Data were obtained from each electrode over a minimum of 8 experimental blocks, each consisting of two presentations (one for each order) of each stimulus pair. The frequency of the ICMS stimuli was 100 Hz and amplitude 60 µA. For ICMS duration, Weber fractions were calculated (as the ratio between just-noticeable-difference and the standard duration) separately for C1 and C2 as we used different duration ranges (stimulus standard was 500ms for C1 and 1s for C2). This task evaluates how well individuals can process and compare temporal variations in sensory stimuli, which is fundamental for interpreting dynamic sensations. To test whether speed discrimination was achieved considering only ICMS train durations, we selected two standards – either motion using PFs at Short Distance (SD, inter-centroids distance of ∼0.6cm) and Long Distance (LD, inter-centroids distance ∼1.4cm) – and paired them with comparison amplitudes that spanned a range of ±500ms around the standard. Tests for both standards were interleaved and randomized within an experimental block in C1. Data were obtained from each combination over a minimum of 8 experimental blocks, each consisting of 2 presentations (one for each order) of each stimulus pair. The frequency of the ICMS stimuli was 100 Hz and amplitude 80 µA. The conversion in actual speed on skin was calculated form the actual distance of the two PFs on skin and the ICMS train duration (Δdistance/Duration). Fitting curves (least-squares) are obtained using *polyfit* function in Matlab. In each speed discrimination task, multiple electrodes were tested on D2 and D3.

### Letter forms encoding and discrimination task

To assess the participant’s ability to make complex perceptual discriminations between different electrical stimulation sequences we used a forced choice discrimination task. Before discrimination testing, C1 drew the perceived pattern to multichannel stimulation several times. We selected from 3 to 6 PFs aligned on D2 in order to create tactile shapes resembling letters (T, L, C, O, I). 500ms ICMS trains were delivered simultaneously from all the selected channels or sequentially from each individual electrode depending on the encoding condition. During each trial of the discrimination task, a single sequence was presented and C1 gave a verbal report to indicate which of the letters the subject had perceived. Sequences were presented in pseudo-random order. Data were obtained over 5 experimental blocks, each consisting of 10 presentations of each stimulus. The frequency of the ICMS stimuli was 100 Hz and amplitude was 80µA. The statistical significance of the accuracy values was obtained using the binom.test function in ‘R’. To calculate complexity, number of bars were presented considering those necessary to draw the specific letter on the skin (**Figure 5C**).

### Psychometric functions

Psychometric functions were fit with a logistic function:

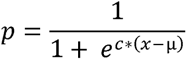

Where *p* is the probability of judging the comparison stimulus as faster than the standard, *x* is the train duration, *c* is the slope, and *µ* the point of subjective equality (PSE). The just noticeable difference (JND) is half the difference between the amplitudes that yield a *p* of 0.25 and 0.75.

### Cortical arrangements

To depict the arrangement of the electrodes adopted in each task, we reported anatomical MRIs with arrays superimposed whose location was based on intra- operative photos (both for C1 and C2) scaled identically. The distances on the cortex (inter-electrode distances) and those on skin (inter-centroids PF distances) were calculated and compared. Pearson’s Correlation of the perceived and cortical edge length (normalized data) was measured. In the motion encoding, arrows indicate the directions of motion on skin and the sequential order of activation of the adopted electrodes on the arrays (lateral and medial).

## Extended Data Figures

**Extended Data Figure 1.**
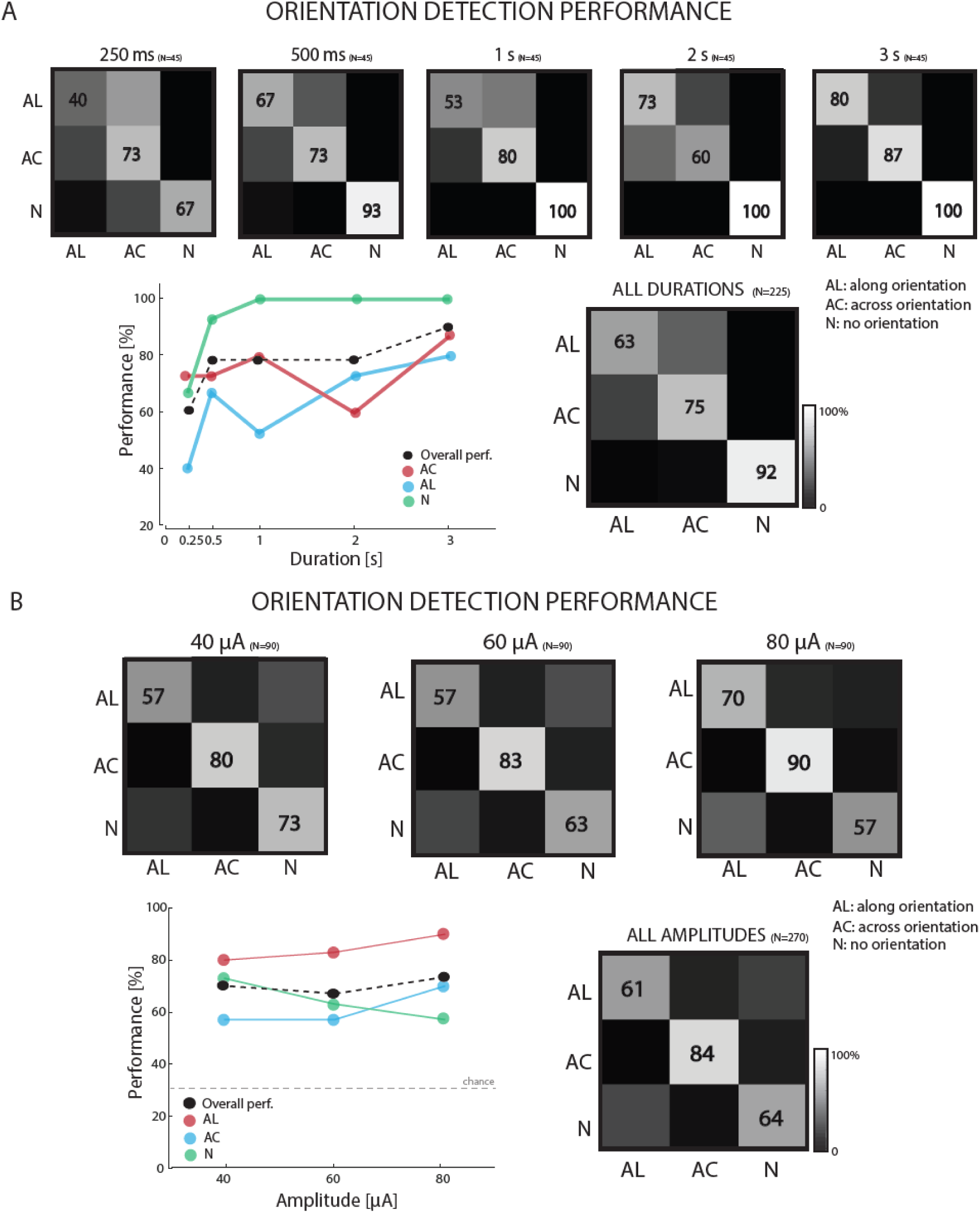
Effects of ICMS train duration and amplitude on orientation recognition. **A|** Orientation recognition performance broken down by ICMS train durations (250ms, 500ms, 1s, 2s, 3s). AC: across the digit, AL: along the length of the digit, N: no orientation (random pattern). N=225. **B|** Orientation recognition performance broken down by ICMS amplitude (40, 60, 80 µA). N=270. Numbers in the boxes indicate the percent of success identifications per condition. Data from C1.

**Extended Data Figure 2.**
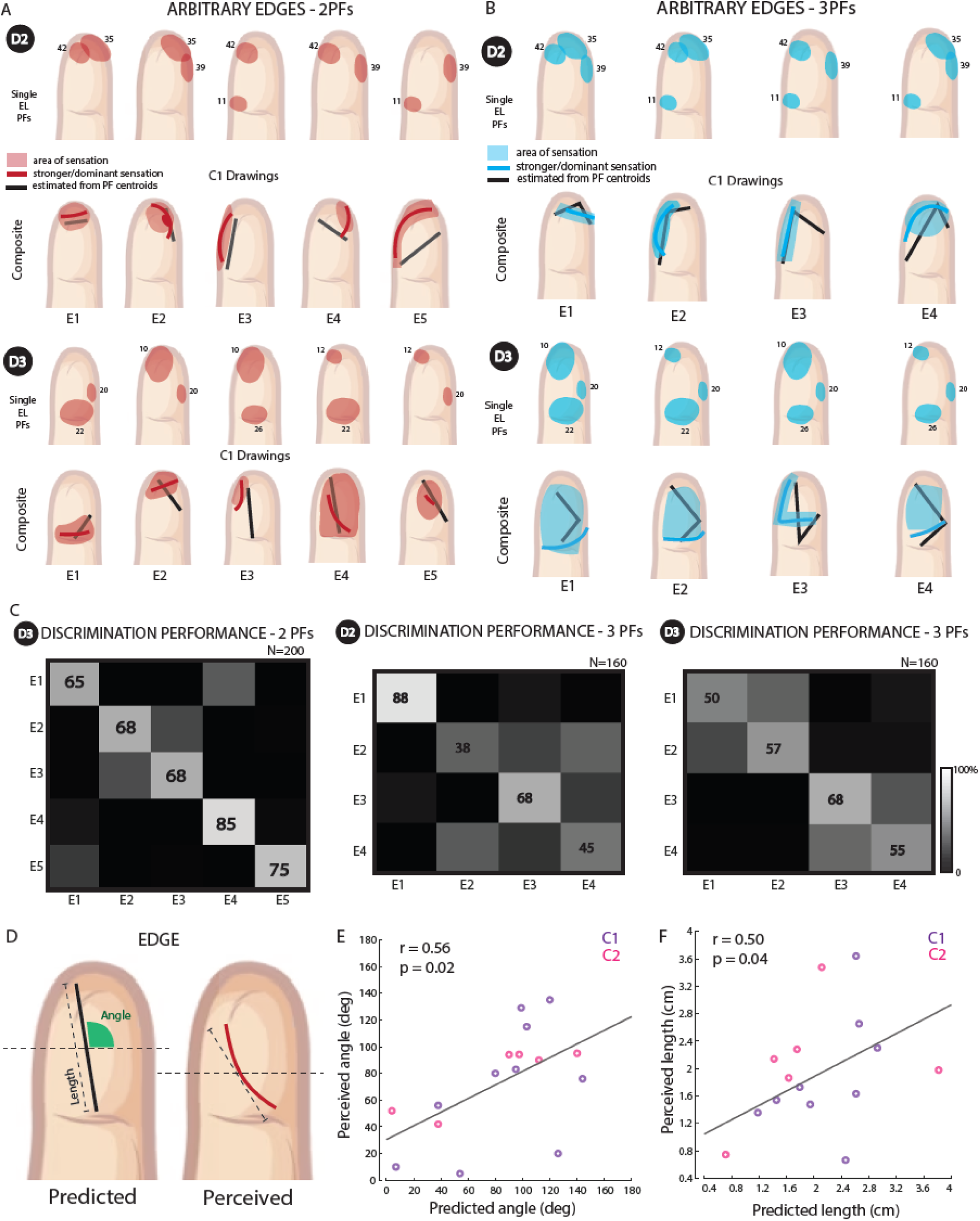
Arbitrary tactile shapes in C1. **A|** All combinations of 2 single electrode PFs to create arbitrary tactile shapes on D2 and D3. Composite percepts reported by C1 when stimulated with the relative combination. **B|** All combinations of 3 single electrode PFs to create arbitrary tactile shapes on D2 and D3. Composite percepts reported by C1 when stimulated with the relative combination. Shaded area represents the area of sensation. Thick lines indicate the zone where the sensation is stronger/more intense. Black lines are the predictions of the evoked sensation from the single electrode PFs. Data from C1. **C|** Identification performance of five arbitrary tactile shapes, combining 2 PFs on D3 (left), randomly presented to C1. Identification performance of four arbitrary tactile shapes, combining 3 PFs on D2 (center) and D3 (right), randomly presented to C1. Numbers in the boxes indicate the percent of success identifications per condition. N=160. **D|** Schematic of predicted and perceived ICMS-evoked sensation angle and length. **E|** Pearson’s Correlation between predicted and perceived sensation angles in both C1 and C2. **F|** Pearson’s Correlation between predicted and perceived sensation lengths in both C1 and C2. Shaded area represents the area of sensation.

**Extended Data Figure 3.**
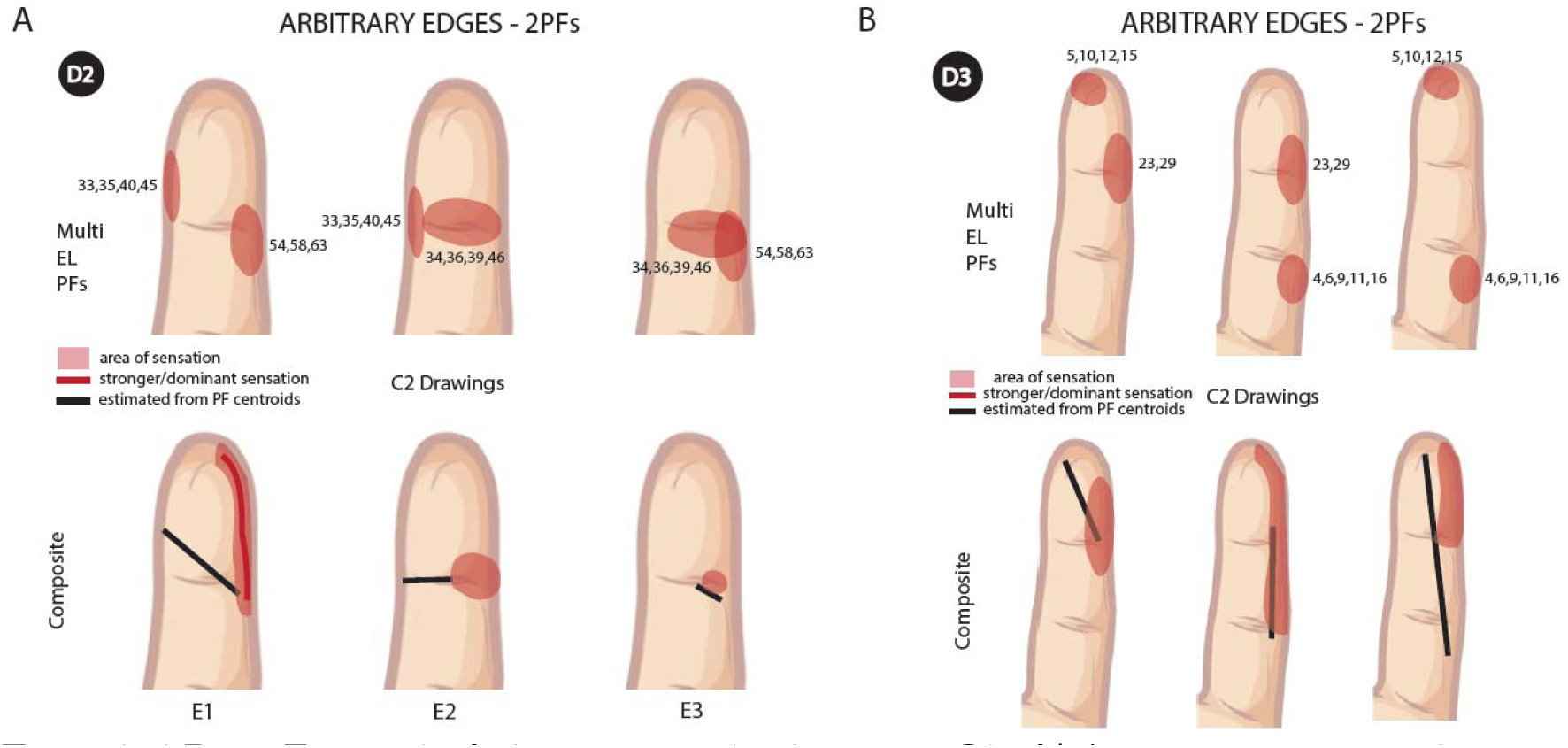
Arbitrary tactile shapes in C2. **A|** All combinations of 2 single electrode PFs to create arbitrary tactile shapes on D2. Composite percepts reported by C1 when stimulated with the relative combination. **B|** All combinations of 2 single electrode PFs to create arbitrary tactile shapes on D3. Composite percepts reported by C1 when stimulated with the relative combination. Data from C2. Shaded area represents the area of sensation. Thick lines indicate the zone where the sensation is stronger/more intense. Black lines are the predictions of the evoked sensation from the single electrode PFs.

**Extended Data Figure 4.**
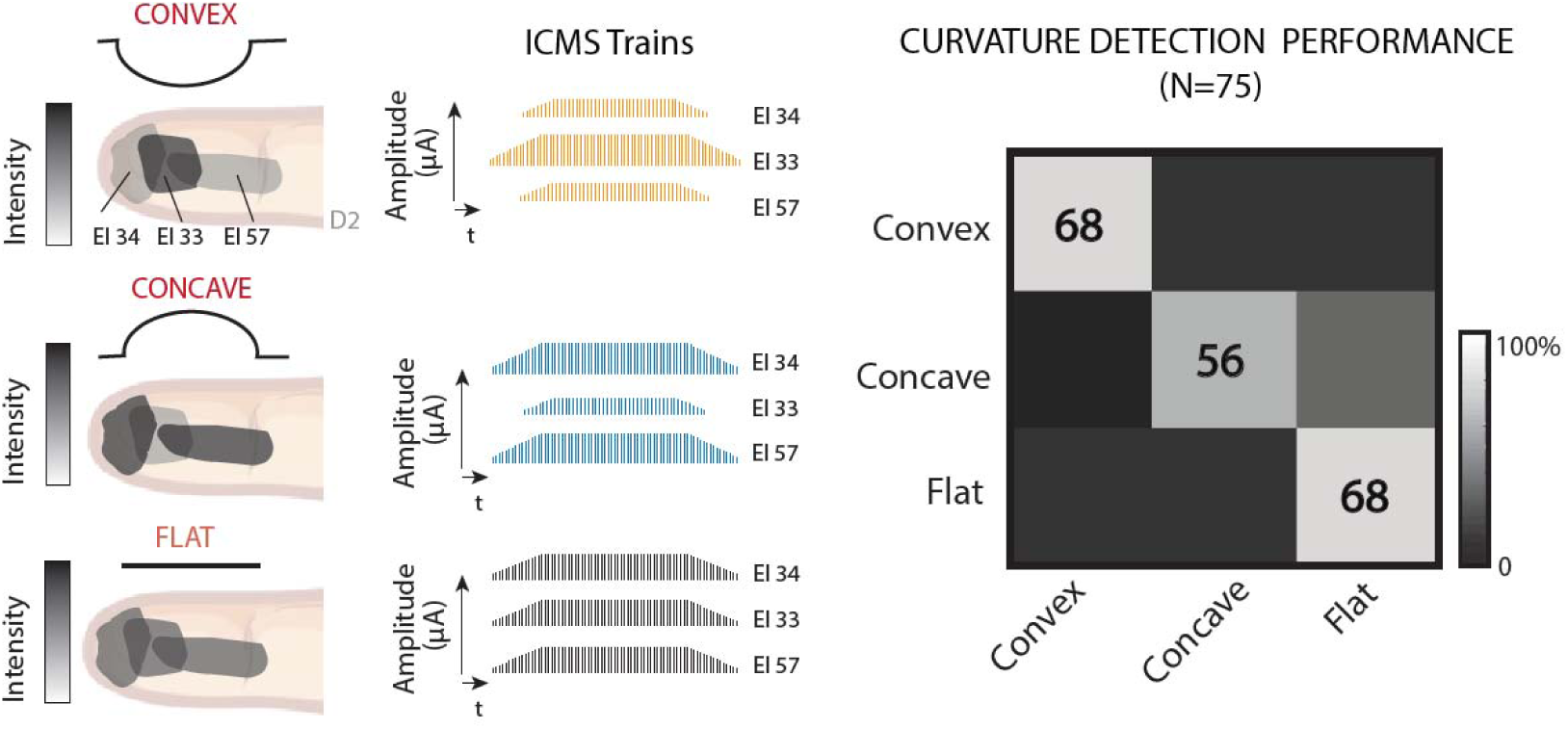
Curvature recognition performance in C1. Curvature recognition performance with 3 different stimuli: convex, concave, and flat edges. N=75. Data from C1.

**Extended Data Figure 5.**
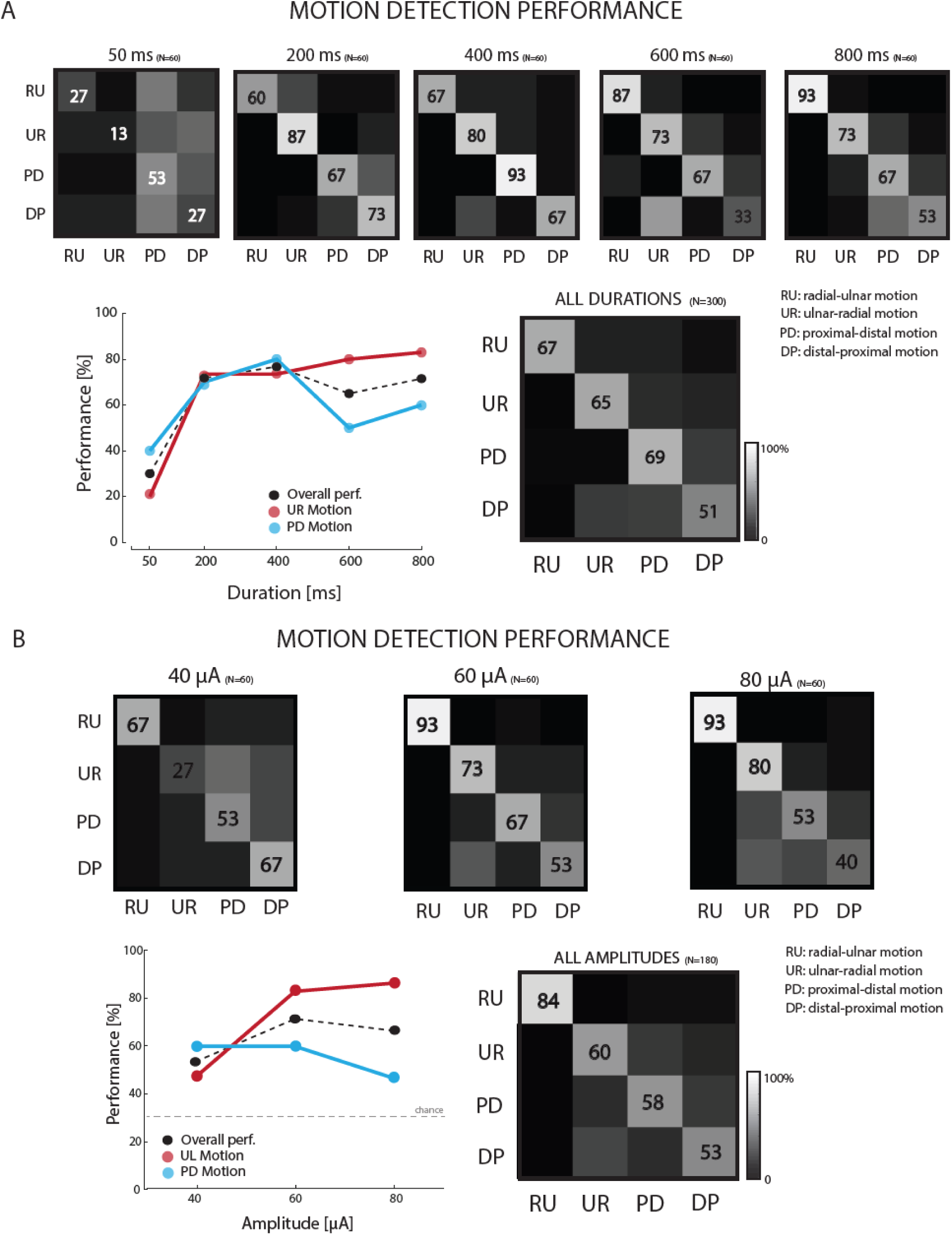
Effects of ICMS duration and amplitude on direction-of- motion recognition. **A|** Direction-of-motion recognition performance broken down by ICMS train durations (50, 200, 400, 600, 800ms). RU: radial-ulnar motion, UR: ulnar-radial motion, PD: proximal-distal motion, DP: distal-proximal motion. N=300. **B|** Direction-of-motion recognition performance broken down by ICMS amplitude (40, 60, 80 µA). N=180. Numbers in the boxes indicate the percent of successful identifications per condition. Data from C1.

**Extended Data Figure 6.**
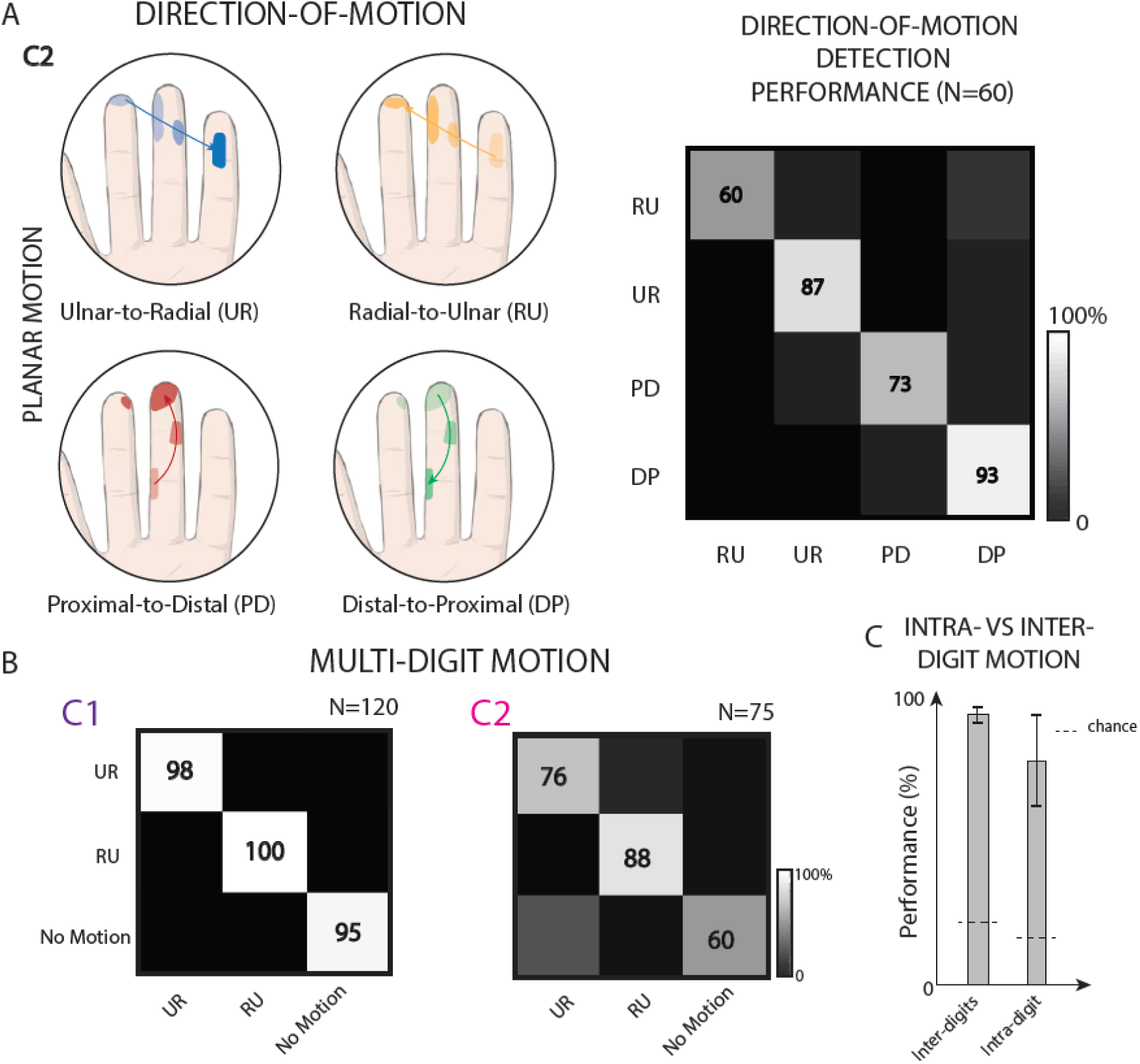
Single and Multi-digit apparent motion. **A|** Combinations of PFs sequentially activated in four different directions (Ulnar-to-Radial, Radial-to-Ulnar, Proximal-to-Distal and Distal-to-Proximal). PFs relate to multi-channel stimulation. Direction-of-motion recognition performance. Numbers in the boxes indicate the percent of success identifications per condition. N=60. Data from C2. **B|** Multidigit motion recognition performance broken down per condition in C1 (N=120) and C2 (N=75). In C1, the apparent motion is across D1, D2, D3, D4 PFs. In C2, it is across D2, D3, D4 PFs. Numbers in the boxes indicate the percent of success identifications per condition. **C|** Comparison of direction-of-motion identification performance between intra- and inter-digits motion.

**Extended Data Figure 7.**
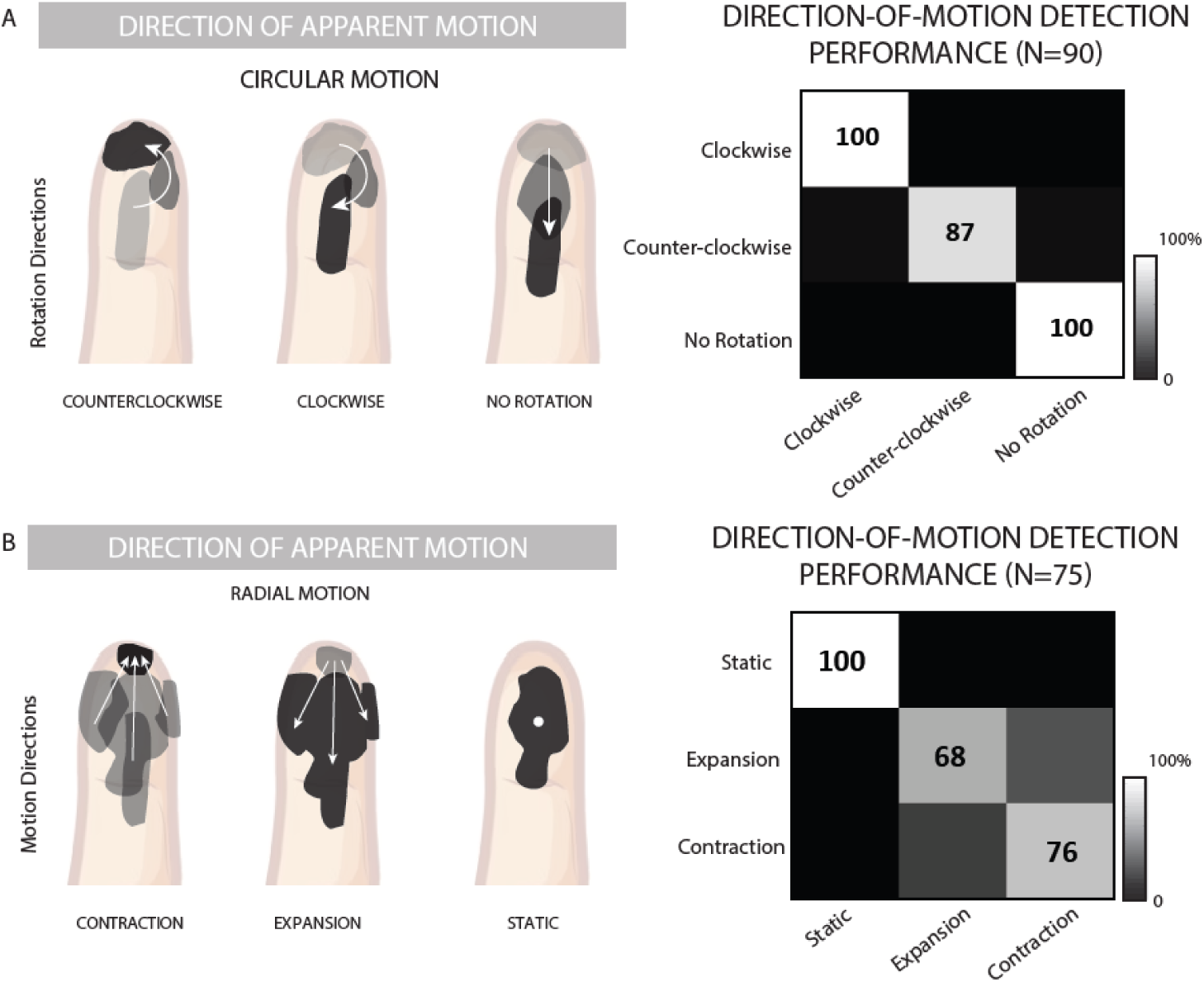
Direction of circular and radial motion recognition. **A|** Combinations of PFs sequentially activated in 3 different directions (Clockwise, Counterclockwise and no Rotation/Rectilinear motion). PFs relate to single channel stimulation. Direction-of-motion recognition performance. N=90. **B|** Combinations of PFs sequentially activated on 3 different directions (Contraction, Expansion and Static). PFs relate to single- or multi-channel stimulation. Direction-of-motion recognition performance. N=75. Numbers in the boxes indicate the percent of success identifications per condition. Data from C1.

**Extended Data Figure 8.**
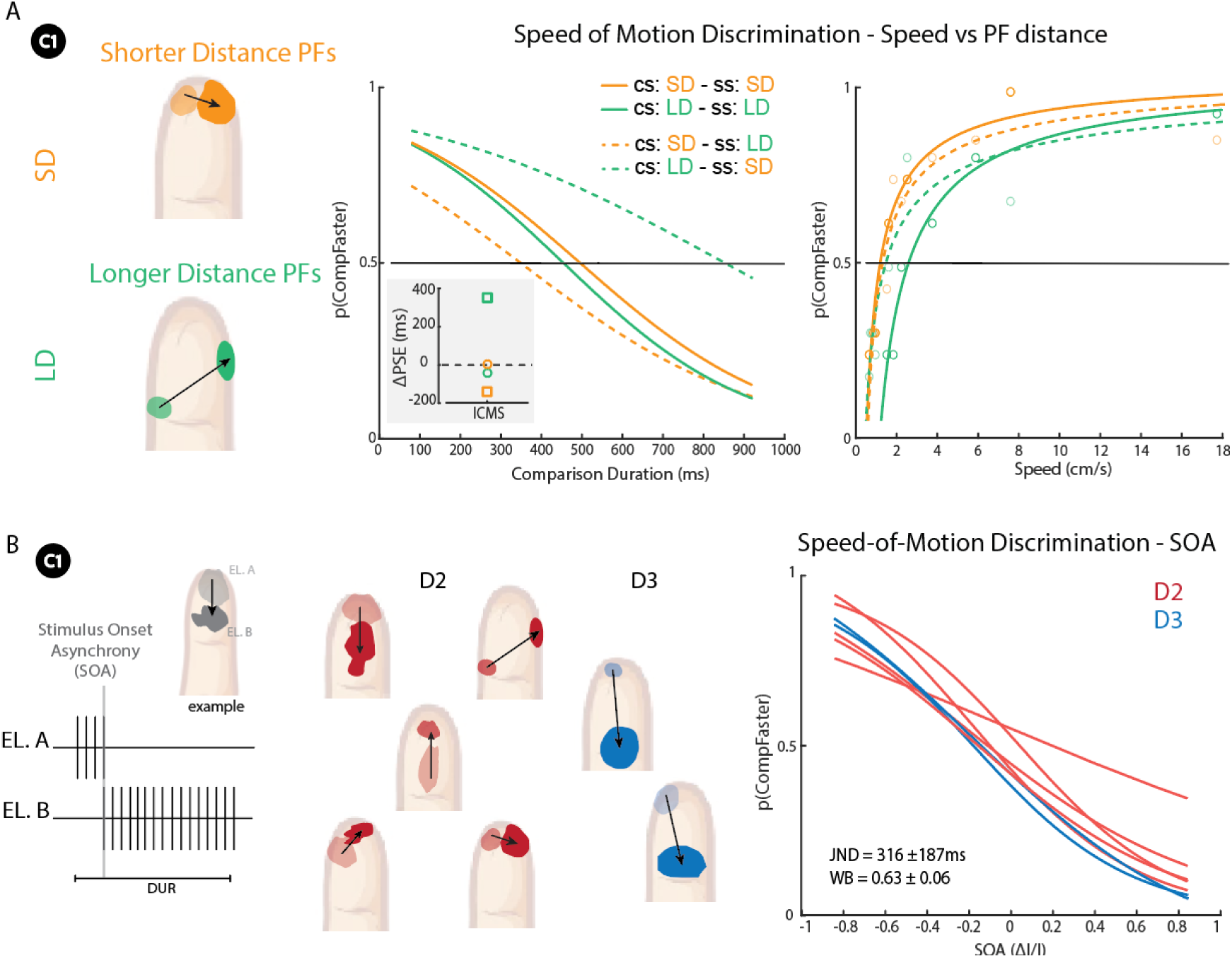
Modulating the speed of motion via ICMS. Encoding motion speed through the modulation of the ICMS train duration between electrodes with different PFs. Short trains encode faster movements on the skin rather than longer ones. **A|** Discrimination task where standard and comparison stimuli are interchanged and delivered through channels evoking PFs at longer (LD) or shorter (SD) distance on D2. Psychometric functions for SD and LD having the same standard (SS) and comparison (SS) (filled lines) or different ones (dashed lines). Inset: variation of PSE when CS and SS are not the same. Curves reported in the speed space considering both distance on skin (cm) and ICMS train duration (ms). **B|** Encoding motion speed through the modulation of the Stimulus Onset Asynchrony (SOA) between electrodes with different PFs. Short SOAs encode faster movements on the skin rather than longer ones. Speed discrimination performance for C1 on both D2 and D3 are reported. Each line corresponds to a 2-electrode combination.

**Extended Data Figure 9.**
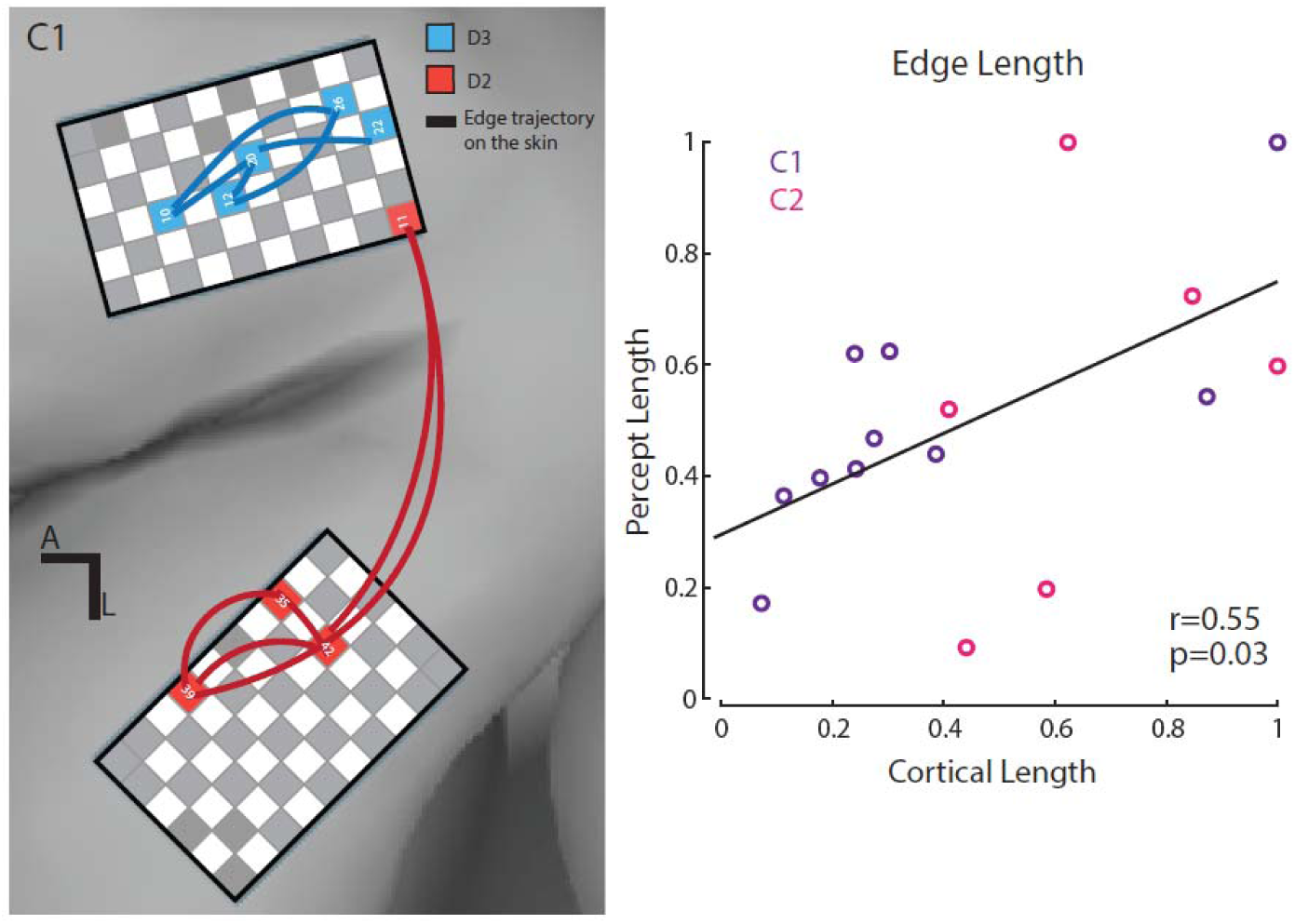
Percept versus Cortical length. Anatomical MRI with arrays superimposed whose location is based on intra-operative photos. L: Lateral. A: Anterior. Cortical distance and ICMS-evoked percepts length. Normalized values with respect to the max distances are reported. Colors and numbers correspond to the electrodes and digit (D2, D3) adopted to encode different stimulus orientations. Pearson’s Correlation between cortical length (distance intra-electrodes) and percept length (drawn by the participant).

**Extended Data Figure 10.**
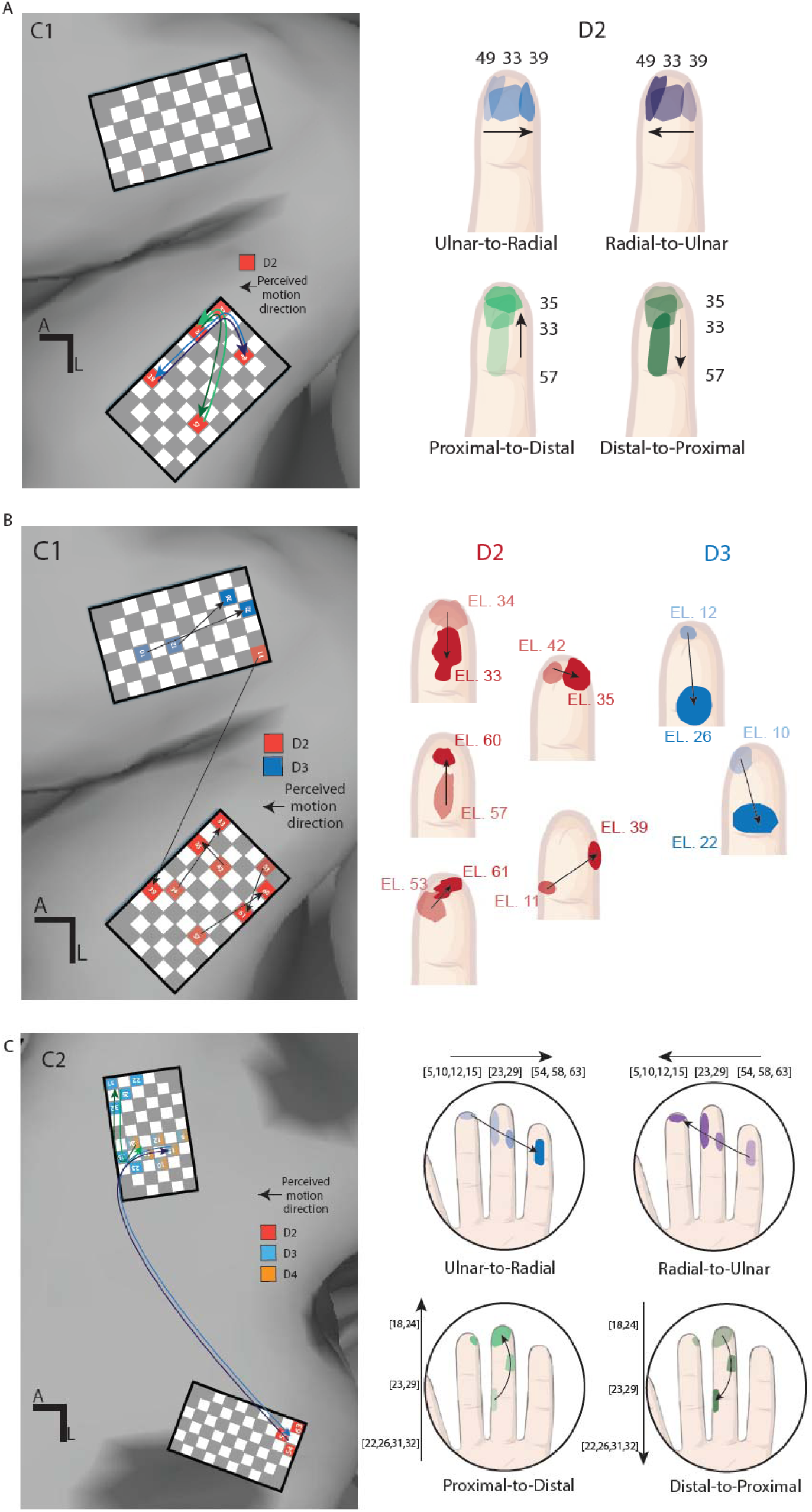
Cortical electrode arrangement. **A|** Cortical directions of apparent motions in C1. Colors and numbers correspond to the electrodes and digits (D2, D3) adopted to encode different directions of motion. Three aligned PFs sequentially activated on D2. PFs adopted in the direction-of-motion detection task. **B|** Two aligned PFs sequentially activated. PFs adopted in the speed discrimination task. Data from C1. **C|** Cortical directions of apparent motions in C2. Anatomical MRI with arrays superimposed whose location is based on intra-operative photos. L: Lateral. A: Anterior. Colors and numbers correspond to the electrodes and digits (D2, D3, D4) adopted to encode different directions of motion. PFs (from mICMS) adopted for the direction of motion recognition task. Data from C2.

## References

1. Bensmaia, S. J., Tyler, D. J. & Micera, S. Restoration of sensory information via bionic hands. Nature Biomedical Engineering 1–13 (2020) doi:10.1038/s41551-020-00630-8.

2. Greenspon, C. M., et al. Biomimetic Multi-Channel Microstimulation of Somatosensory Cortex Conveys High Resolution Force Feedback for Bionic Hands. http://biorxiv.org/lookup/doi/10.1101/2023.02.18.528972 (2023) doi:10.1101/2023.02.18.528972.

3. Hughes, C. L. et al. Perception of microstimulation frequency in human somatosensory cortex. eLife 10, e65128 (2021).

4. Pei, Y.-C. & Bensmaia, S. J. The neural basis of tactile motion perception. J Neurophysiol 112, 3023–3032 (2014).

5. Pack, C. C. & Bensmaia, S. J. Seeing and Feeling Motion: Canonical Computations in Vision and Touch. PLoS Biol 13, e1002271 (2015).

6. Johansson, R. S. & Flanagan, J. R. Coding and use of tactile signals from the fingertips in object manipulation tasks. Nature Reviews Neuroscience 10, 345–359 (2009).

7. Kim, S. S., Gomez-Ramirez, M., Thakur, P. H. & Hsiao, S. S. Multimodal Interactions between Proprioceptive and Cutaneous Signals in Primary Somatosensory Cortex. Neuron 86, 555–566 (2015).

8. Pei, Y.-C., Hsiao, S. S., Craig, J. C. & Bensmaia, S. J. Neural Mechanisms of Tactile Motion Integration in Somatosensory Cortex. Neuron 69, 536–547 (2011).

9. Collinger, J. L. et al. Functional Priorities, Assistive Technology, and Brain-Computer Interfaces after Spinal Cord Injury. J Rehabil Res Dev 50, 145–160 (2013).

10. Wodlinger, B. et al. Ten-dimensional anthropomorphic arm control in a human brain−machine interface: difficulties, solutions, and limitations. J. Neural Eng. 12, 016011 (2014).

11. Flesher, S. N. et al. Intracortical microstimulation of human somatosensory cortex. Science Translational Medicine 8, 361ra141–361ra141 (2016).

12. Salas, M. A. et al. Proprioceptive and cutaneous sensations in humans elicited by intracortical microstimulation. eLife 7, 1–11 (2018).

13. Fifer, M. S. et al. Intracortical Somatosensory Stimulation to Elicit Fingertip Sensations in an Individual With Spinal Cord Injury. Neurology 98, e679–e687 (2022).

14. Greenspon, C. M. et al. Tessellation Of Artificial Touch Via Microstimulation Of Human Somatosensory Cortex. 2023.06.23.545425 Preprint at 10.1101/2023.06.23.545425 (2023).

15. Bjånes, D. A. et al. Multi-channel intra-cortical micro-stimulation yields quick reaction times and evokes natural somatosensations in a human participant. medRxiv 2022.08.08.22278389 (2022) doi:10.1101/2022.08.08.22278389.

16. Flesher, S. N. et al. A brain-computer interface that evokes tactile sensations improves robotic arm control. Science 372, 831–836 (2021).

17. Pruszynski, J. A., Flanagan, J. R. & Johansson, R. S. Fast and accurate edge orientation processing during object manipulation. eLife 7, e31200.

18. Cho, Y., Craig, J. C., Hsiao, S. S. & Bensmaia, S. J. Vision is superior to touch in shape perception even with equivalent peripheral input. Journal of Neurophysiology 115, 92–99 (2016).

19. Bensmaia, S. J., Hsiao, S. S., Denchev, P. V., Killebrew, J. H. & Craig, J. C. The tactile perception of stimulus orientation. Somatosensory & Motor Research 25, 49–59 (2008).

20. Renfrew, S. & Cavanagh, D. THE DISCRIMINATION BETWEEN PINCHING AND PRESSING OF THE SKIN: THE BASIS OF A CLINICAL TEST. Brain 77, 305–313 (1954).

21. Gardner, E. P. & Costanzo, R. M. Spatial integration of multiple-point stimuli in primary somatosensory cortical receptive fields of alert monkeys. Journal of Neurophysiology 43, 420–443 (1980).

22. Chen, L. M., Friedman, R. M. & Roe, A. W. Optical imaging of a tactile illusion in area 3b of the primary somatosensory cortex. Science 302, 881–885 (2003).

23. Yau, J. M., Connor, C. E. & Hsiao, S. S. Representation of tactile curvature in macaque somatosensory area 2. Journal of Neurophysiology 109, 2999–3012 (2013).

24. Phillips, J. R. & Johnson, K. O. Tactile spatial resolution. II. Neural representation of Bars, edges, and gratings in monkey primary afferents. J Neurophysiol 46, 1192–1203 (1981).

25. Goodwin, A. W., John, K. T. & Marceglia, A. H. Tactile discrimination of curvature by humans using only cutaneous information from the fingerpads. Exp Brain Res 86, 663–672 (1991).

26. Kappers, A. M. L. Human perception of shape from touch. Philos Trans R Soc Lond B Biol Sci 366, 3106– 3114 (2011).

27. Hsiao, S. Central mechanisms of tactile shape perception. Current Opinion in Neurobiology 18, 418–424 (2008).

28. Suresh, A. K., Saal, H. P. & Bensmaia, S. J. Edge orientation signals in tactile afferents of macaques. Journal of Neurophysiology 116, 2647–2655 (2016).

29. Leslie L. Clark. Research Bulletin. (American Foundation for the Blind, 1965).

30. Kirman, J. H. Tactile apparent movement: The effects of interstimulus onset interval and stimulus duration. Perception & Psychophysics 15, 1–6 (1974).

31. Sherrick, C. E. & Rogers, R. Apparent haptic movement. Perception & Psychophysics 1, 175–180 (1966).

32. Kolers, P. A. The illusion of movement. Scientific American 211, 98–106 (1964).

33. Gardner, E. P. Somatosensory cortical mechanisms of feature detection in tactile and kinesthetic discrimination. Can. J. Physiol. Pharmacol. 66, 439–454 (1988).

34. Warren, S., Hamalainen, H. A. & Gardner, E. P. Objective classification of motion- and direction-sensitive neurons in primary somatosensory cortex of awake monkeys. J Neurophysiol 56, 598–622 (1986).

35. Beauchamp, M. S. et al. Dynamic Stimulation of Visual Cortex Produces Form Vision in Sighted and Blind Humans. Cell 181, 774–783.e5 (2020).

36. Firszt, J. B., Koch, D. B., Downing, M. & Litvak, L. Current Steering Creates Additional Pitch Percepts in Adult Cochlear Implant Recipients. Otology & Neurotology 28, 629 (2007).

37. Ogrinc, M., Farkhatdinov, I., Walker, R. & Burdet, E. Sensory integration of apparent motion speed and vibration magnitude. IEEE Trans Haptics 11, 455–463 (2018).

38. Edin, B. B., Essick, G. K., Trulsson, M. & Olsson, K. A. Receptor encoding of moving tactile stimuli in humans. I. Temporal pattern of discharge of individual low-threshold mechanoreceptors. J Neurosci 15, 830–847 (1995).

39. Saal, H. P., Delhaye, B. P., Rayhaun, B. C. & Bensmaia, S. J. Simulating tactile signals from the whole hand with millisecond precision. Proc Natl Acad Sci USA 114, E5693–E5702 (2017).

40. Lashley, K. S. The problem of serial order in behavior. in Cerebral mechanisms in behavior; the Hixon Symposium 112–146 (Wiley, Oxford, England, 1951).

41. Gibson, J. J. Observations on active touch. Psychological Review 69, 477–491 (1962).

42. Valle, G. et al. Biomimetic Intraneural Sensory Feedback Enhances Sensation Naturalness, Tactile Sensitivity, and Manual Dexterity in a Bidirectional Prosthesis. Neuron 100, 37–45.e7 (2018).

43. Valle, G. et al. Biomimetic computer-to-brain communication enhancing naturalistic touch sensations via peripheral nerve stimulation. Nat Commun 15, 1151 (2024).

44. Yau, J. M., Pasupathy, A., Fitzgerald, P. J., Hsiao, S. S. & Connor, C. E. Analogous intermediate shape coding in vision and touch. Proceedings of the National Academy of Sciences 106, 16457–16462 (2009).

45. Kumaravelu, K., Sombeck, J., Miller, L. E., Bensmaia, S. J. & Grill, W. M. Stoney vs. Histed: Quantifying the spatial effects of intracortical microstimulation. Brain Stimulation 15, 141–151 (2022).

46. Beyeler, M., Rokem, A., Boynton, G. M. & Fine, I. Learning to see again: biological constraints on cortical plasticity and the implications for sight restoration technologies. J Neural Eng 14, 051003 (2017).

47. Chen, X., Wang, F., Fernandez, E. & Roelfsema, P. R. Shape perception via a high-channel-count neuroprosthesis in monkey visual cortex. Science 370, 1191–1196 (2020).

48. Fernández, E. et al. Visual percepts evoked with an intracortical 96-channel microelectrode array inserted in human occipital cortex. J Clin Invest 131, (2021).

49. Oswalt, D. et al. Multi-electrode stimulation evokes consistent spatial patterns of phosphenes and improves phosphene mapping in blind subjects. Brain Stimulation 14, 1356–1372 (2021).

50. Kumaravelu, K. & Grill, W. M. Neural mechanisms of the temporal response of cortical neurons to intracortical microstimulation. Brain Stimul 17, 365–381 (2024).

51. Penfield, W. & Boldrey, E. SOMATIC MOTOR AND SENSORY REPRESENTATION IN THE CEREBRAL CORTEX OF MAN AS STUDIED BY ELECTRICAL STIMULATION1. Brain 60, 389– 443 (1937).

52. Yau, J. M., Kim, S. S., Thakur, P. H. & Bensmaia, S. J. Feeling form: the neural basis of haptic shape perception. Journal of Neurophysiology 115, 631–642 (2016).

53. Valle, G. et al. Comparison of linear frequency and amplitude modulation for intraneural sensory feedback in bidirectional hand prostheses. Scientific Reports 8, 16666 (2018).

54. D’Anna, E. et al. A closed-loop hand prosthesis with simultaneous intraneural tactile and position feedback | Science Robotics. Science Robotics 4, (2019).

55. Osborn, L. E. et al. Intracortical microstimulation of somatosensory cortex enables object identification through perceived sensations. in 2021 43rd annual international conference of the IEEE engineering in medicine & biology society (EMBC) 6259–6262 (IEEE, 2021).

56. Zaaimi, B., Ruiz-Torres, R., Solla, S. A. & Miller, L. E. Multi-electrode stimulation in somatosensory cortex increases probability of detection. J. Neural Eng. 10, 056013 (2013).

57. Shelchkova, N. D. et al. Microstimulation of human somatosensory cortex evokes task-dependent, spatially patterned responses in motor cortex. Nat Commun 14, 7270 (2023).

58. Donati, E. & Valle, G. Neuromorphic hardware for somatosensory neuroprostheses. Nat Commun 15, 556 (2024).

59. Long, K. H., Lieber, J. D. & Bensmaia, S. J. Texture is encoded in precise temporal spiking patterns in primate somatosensory cortex. Nat Commun 13, 1311 (2022).

60. Chortos, A., Liu, J. & Bao, Z. Pursuing prosthetic electronic skin. Nat Mater 15, 937–950 (2016).

61. Wang, W. et al. Neuromorphic sensorimotor loop embodied by monolithically integrated, low-voltage, soft e-skin. Science 380, 735–742 (2023).

