## Supplementary Information for "TACTILE EDGES AND MOTION VIA PATTERNED MICROSTIMULATION OF THE HUMAN CORTEX"

**List of Supplementary Materials:**

Movies S1: Tactile edge encoding, C1 and C2.

Movies S2: Tactile curvature encoding, C1.

Movies S3: 3D structures from multidigit tactile edges, C1.

Movies S4: Apparent motion encoding, C1 and C2.

Movies S5: Dynamic voltage field shaping, C1.

Movies S6: Speed of motion encoding, C1.
